# Evolutionary dynamics of 1,976 lymphoid malignancies predict clinical outcome

**DOI:** 10.1101/2023.11.10.23298336

**Authors:** Calum Gabbutt, Martí Duran-Ferrer, Heather Grant, Diego Mallo, Ferran Nadeu, Jacob Househam, Neus Villamor, Olga Krali, Jessica Nordlund, Thorsten Zenz, Elias Campo, Armando Lopez-Guillermo, Jude Fitzgibbon, Chris P Barnes, Darryl Shibata, José I Martin-Subero, Trevor A Graham

**Affiliations:** Centre for Evolution and Cancer, Institute of Cancer Research, London, UK; Centre for Genomics and Computational Biology, Barts Cancer Institute, Queen Mary University of London, London, UK; Fundació de Recerca Clínic Barcelona-Institut d’Investigacions Biomèdiques August Pi i Sunyer (FRCB-IDIBAPS), Barcelona, Spain; Arizona Cancer Evolution Center, Biodesign Institute and School of Life Sciences, Arizona State University, Tempe, AZ, United States; Centro de Investigación Biomédica en Red de Cáncer (CIBERONC), Madrid, Spain; Hospital Clínic de Barcelona, Barcelona, Spain; Department of Medical Sciences, Molecular Precision Medicine and Science for Life Laboratory, Uppsala University, Uppsala, Sweden; Department of Medical Oncology and Hematology, University Hospital and University of Zürich, Zurich; Departament de Fonaments Clínics, Universitat de Barcelona, Barcelona, Spain; Department of Cell & Developmental Biology, University College London, London, UK; University of Southern California, Keck School of Medicine, Department of Pathology, Los Angeles, CA, USA; Institució Catalana de Recerca i Estudis Avançats (ICREA), Barcelona, Spain

## Abstract

Cancer development, progression, and response to treatment are evolutionary processes, but characterising the evolutionary dynamics at sufficient scale to be clinically-meaningful has remained challenging. Here, we develop a new methodology called EVOFLUx, based upon natural DNA methylation barcodes fluctuating over time, that quantitatively infers evolutionary dynamics using only a bulk tumour methylation profile as input. We apply EVOFLUx to 1,976 well-characterised lymphoid cancer samples spanning a broad spectrum of diseases and show that tumour growth rates, malignancy age and epimutation rates vary by orders of magnitude across disease types. We measure that subclonal selection occurs only infrequently within bulk samples and detect occasional examples of multiple independent primary tumours. Clinically, we observe that tumour growth rates are higher in more aggressive disease subtypes, and in two series of chronic lymphocytic leukaemia patients, evolutionary histories are independent prognostic factors. Phylogenetic analyses of longitudinal CLL samples using EVOFLUx detect the seeds of future Richter transformation many decades prior to presentation. We provide orthogonal verification of EVOFLUx inferences using additional genetic and clinical data. Collectively, we show how widely-available, low-cost bulk DNA methylation data precisely measures cancer evolutionary dynamics, and provides new insights into cancer biology and clinical behaviour.

## Introduction

The growth and dissemination of cancer is fundamentally an evolutionary process (Nowell 1976; Merlo et al. 2006); consequently, the past evolutionary trajectory of a cancer should strongly influence its future trajectory and allow inference of the clinical path of a patient (Greaves and Maley 2012). However, testing this hypothesis directly is challenging as a precise characterization of cancer evolutionary dynamics would require the analysis of multiple sequential patient samples. Instead, evolutionary trajectories must be inferred by means of a proxy; for example, somatic (epi)mutations are patterned in distinctive ways by differing evolutionary dynamics (Turajlic et al. 2019). In the haematological system, genome sequencing of single cells or single cell colonies are employed to infer the phylogenetic relationships among cells (Gaiti et al. 2019; Lee-Six et al. 2018; N. Williams et al. 2022). The expense of this approach has restricted analyses to small numbers of cases, which limits its scaling for clinical translation.

DNA methylation can also be employed as a heritable lineage marker, recording the clonal architecture of a population of cells (Yatabe, Tavaré, and Shibata 2001; Hong et al. 2010; Brocks et al. 2014; Hao et al. 2016; Siegmund et al. 2009) or the total cell proliferative history (Duran-Ferrer et al. 2020; Endicott et al. 2022). We have recently identified CpGs whose methylation state in each allele is heritable, but stochastically fluctuate over time. These fluctuating CpGs (fCpGs) function as a methylation barcode and so offer a low-cost strategy to provide high temporal resolution lineage tracing in patient samples (Gabbutt et al. 2022).

In a single diploid region, an fCpG can take one of three states: neither allele methylated (0% methylated), one allele methylated (50% methylated) or both alleles methylated (100% methylated). If multiple fCpGs are considered together, collectively they represent a vast number of possibilities (i.e. 3 to the power of the number of fCpGs) where distantly related cells carry distinct fCpG barcodes. These barcodes are not stable, but stochastically methylate and demethylate over time at a timescale measured in years (Gabbutt et al. 2022). Therefore, they not only provide a means to trace the ancestor of a clonal expansion, but also an “evolving barcode” that tracks clonal evolution: two somatic cells with close ancestry will share a near-identical pattern of fCpG methylation, whereas distantly related cells will have divergent fCpG methylation patterns. In bulk populations of somatic cells, the dominant fCpG pattern in the sample represents the fCpG state of the founder cell of the population and intermediate methylation values that deviate from 0, 0.5 and 1 are caused by subclonal expansions in the population (fig 1A). Consequently, the full bulk distribution of fCpG methylation is determined by the evolution of the population and so provides a precise proxy-measurement of the evolutionary dynamics.

Here, we use fCpGs to infer the evolutionary dynamics of cancer cells from clinical specimens, at scale. We focus on lymphoid neoplasms, which cover a broad spectrum of diseases and subtypes with highly variable biological features and clinical manifestations (de Leval et al. 2022; Arber et al. 2022). These tumours have been extensively profiled by DNA methylation arrays, which have provided important insights into the cellular origin, pathogenesis, and clinical behaviour of these tumours (Oakes and Martin-Subero 2018). While their temporal clonal dynamics has been analysed in part (Gruber et al. 2019; Gutierrez et al. 2021), their precise evolutionary dynamics remains poorly characterised.

We construct a quantitative modelling framework called “EVOlutionary inference using FLUctuating methylation” (EVOFLUx) that enables precise quantitative estimation of cancer cell dynamics in lymphoid tumours from individual patients. In 1,976 individual lymphoid malignancies, we precisely measure evolutionary history and show that the patient-specific evolutionary dynamics is strongly associated with disease outcomes.

## Results

### Bulk Methylation Array Samples from 2,430 samples

We assembled and processed with a harmonized pipeline 2,430 bulk sample Illumina methylation array data of normal and neoplastic lymphoid cells (Kulis et al. 2015; Nordlund et al. 2013; Reinius et al. 2012; S. T. Lee et al. 2015; Queirós et al. 2016; Nadeu et al. 2020; Duran-Ferrer et al. 2020; Nadeu et al. 2022; Oakes et al. 2016; Dietrich et al. 2018; Agirre et al. 2015). (Methods). After quality control, we retained 2,204 samples from 2,054 patients (including 22 technical replicates, 3 synchronic and 125 longitudinal samples from the same patients; tables 1, S1-2) and 389,180 CpGs for downstream analyses. As healthy control samples, this dataset contained sorted CD19^+^ B cells (n=40), CD3^+^ T cells (n=35), peripheral blood mononuclear cells (PBMCs, n=6) and whole blood samples (n=6). As tumor samples, we included precursor 797 B- and 90 T-acute lymphoblastic leukemias (B- and T-ALL, respectively) at diagnosis, 28 B and 2 T-ALL at relapse as well as 74 B and 12 T-ALL at complete remission (i.e. normal blood); 149 mantle cell lymphomas (MCL); 722 chronic lymphocytic leukemias (CLL), 55 of its precursor condition monoclonal B-cell lymphocytosis (MBL), and 6 samples from CLL patients undergoing a diffuse large B-cell lymphoma (DLBCL) transformation called Richter transformation (RT); 62 primary DLBCL, not otherwise specified (NOS); and 104 multiple myeloma (MM) and 16 of its precursor condition monoclonal gammopathy of undetermined significance (MGUS). In addition, clinical follow-up was available for 1,667 patients and matched DNA methylation with genetic (whole genome/exome sequencing (WGS/WES)) or with RNA-seq data was available for 505 and 293 CLL/RT samples, respectively (tables 1, S1). Therefore, this exquisite dataset was unique from multiple perspectives, including tumors derived from lymphocytes at various maturation stages, tumors with a highly proliferative acute presentation and more indolent chronic leukemias, tumors from patients ranging from infants to old adults, and tumor samples from different niches and disease stages with available genetic, transcriptomic, and clinical data.

**Table 1:**
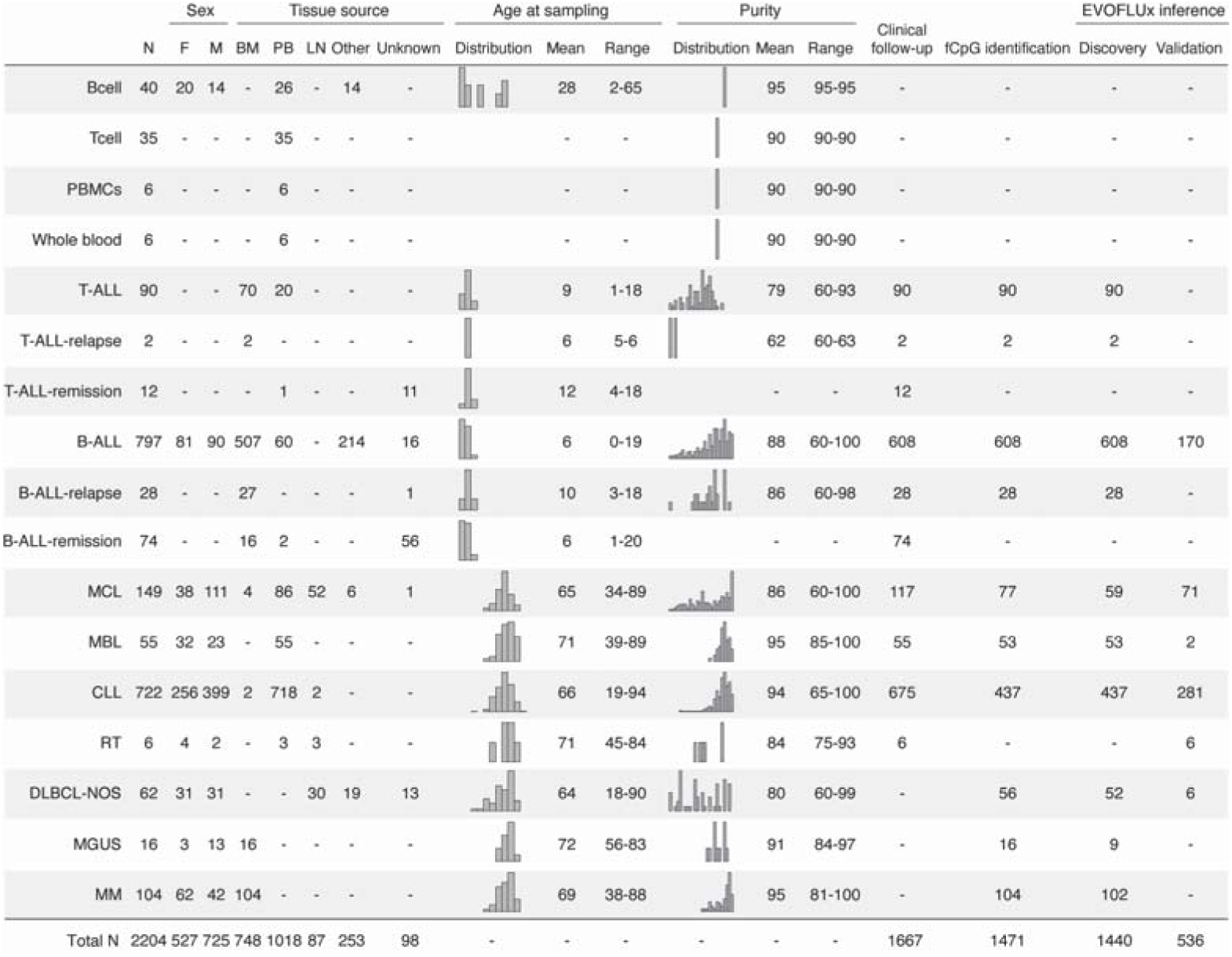
A summary of methylation array samples. Abbreviations: Peripheral blood mononuclear cells (PBMCs); precursor T-acute lymphoblastic leukemias (T-ALL); precursor B-acute lymphoblastic leukemias (B-ALL); mantle cell lymphoma; monoclonal B-cell lymphocytosis (MBL); chronic lymphocytic leukemias (CLL); Richter transformation (RT); diffuse large B-cell lymphoma, not otherwise specified (DLBCL-NOS); monoclonal gammopathy of undetermined significance (MGUS); and multiple myeloma (MM). Female (F); Male (M); bone marrow (BM); peripheral blood (PB); lymph node (LN).

### Identification of fCpG Loci in Blood Malignancies

We have previously observed that fCpG loci are tissue-specific (Gabbutt et al. 2022). To identify fCpGs in the lymphoid system we first partitioned our dataset into discovery (n=1,471) and validation sets (n=733, tumour samples n=536; tables 1, S2). Using the discovery set, we selected for CpG loci that fulfilled the following criteria:

i. Heterogeneous across different subjects with the same disease (by accepting CpG loci with top 5% of standard deviation of methylation value within a cancer type)
ii. Equally likely to be methylated or unmethylated (by averaging across all cases in a cancer type and accepting only CpG sites with average methylation of approximately 0.5 in the cancer type)
iii. Unlikely to be associated with specific cell or cancer types. This was done using an unsupervised Laplacian Score (LS) feature selection metric (He, Cai, and Niyogi 2005) that ranked CpG loci by their tendency to preserve the nearest-neighbour graph, where we accepted the 5% least informative CpG loci.

To visualise how the LS quantified the informativeness of individual CpG loci, we performed a principal component analysis (PCA) on methylation values (fig. 1B). The first 2 principal components revealed disease specific clustering in methylation. The CpG loci with the most informative features for clustering (lowest LS) showed disease-specific methylation. In contrast, the CpG loci that were least informative for clustering (highest LS) tended to have either low variability or high variability that was well-dispersed across the PCA. Combining these filters yielded 978 pan-lymphoid cancer fCpGs (table S3). These fCpGs did not cluster the samples by either disease (fig. 1C) or disease-subtype in either discovery or validation samples (fig. S1A-D), nor with the version of methylation array employed.

**Figure 1.**
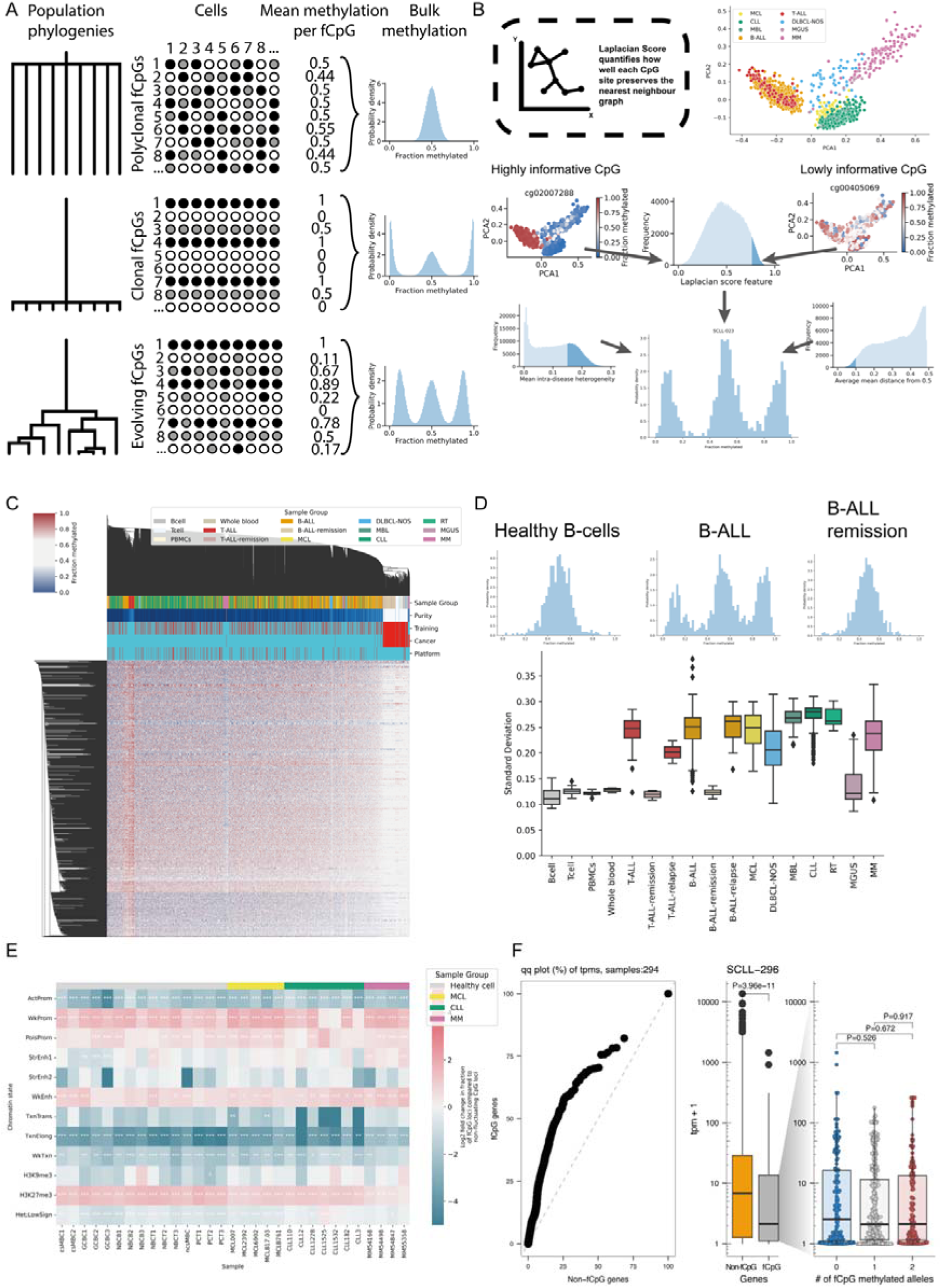
Selection and properties of fCpG loci. **A:** Illustration of how fCpGs enable tracking of lineage dynamics, describing three limiting cases of a population evolutionary structure: (top) polyclonal, in which the most recent common ancestor (MRCA) exists far in the past; (middle) clonal, in which the MRCA is very recent; (bottom) ongoing evolution following a population bottleneck. In a polyclonal population, the different fCpGs (white – unmethylated, grey – heterozygous methylated, black – methylated) are all different due to fluctuations that occurred since the MRCA. Thus, the average methylation of the population as each fCpG is roughly the steady state methylation value. Following a clonal expansion, all the fCpGs in the population are the same, so the average methylation reflects that of the methylation of the founder cell. However, as the population continues to grow ongoing fluctuations alter the barcodes in individual cells from the ancestral barcode, changing the distribution of bulk methylation values. **B:** fCpGs were selected by combining three filters: (i) CpGs with low intra-disease heterogeneity were removed as sites that were likely not fluctuating, (ii) CpGs with a mean methylation far from 0.5 were removed as likely belonging to CpG sites with skewed methylation/demethylation rates, and (iii) CpGs which preserved the nearest-neighbour graph (quantified using the Laplacian Score) were removed as sites that were likely under selection/strict regulation. **C:** A hierarchically-clustered heatmap of the 978 fCpGs. **D:** (top) Example methylation distributions of the fCpGs from healthy B cells, B-ALL cells and whole blood from a patient in remission from B-ALL. (bottom) Boxplots (whiskers extending to ±1.5×IQR) displaying the standard deviation of the methylation distributions separated by cell/disease type. **E:** A heatmap showing the log2 fold change in chromatin status at the site of fCpG loci compared to other CpG loci on the 450K Illumina array in healthy lymphoid cells, MCL, CLL and MM samples. White stars signify the degree of statistical significance (_x2_ tests, hs correction). **F:** Expression analysis on the genes associated with fCpG and non-fCpG loci: (left) a Q-Q comparing non-fCpG and fCpG associated genes, demonstrating fCpG associated genes have a lower expression (in transcripts per million), (middle) a comparison between fCpG and non-fCpG associated genes in a single CLL sample (_P_=3.96e10^-11^, Wilcoxon test), (right) the expression of fCpG associated genes separated by discretised methylation status (all p>0.05, Wilcoxon test).

We then examined the distribution of these 978 fCpG methylation values in individual samples (table S4). As we previously predicted (Gabbutt et al. 2022), in each cancer sample the fCpG loci followed a characteristic “W-shaped” distribution that depicts the fCpG methylation patterns of the founder cell of the cancer sample (fig 1D). In contrast, the healthy B cell subpopulations, which were not included in the discovery set, had unimodal distributions with intermediate methylation consistent with these being polyclonal populations (fig. 1D). The fCpG methylation distributions of samples from ALL patients in remission (i.e. contain mostly healthy cells) similarly had unimodal distributions and clustered with the normal B cell subpopulations (fig 1C-D).

Employing the standard deviation of each sample’s fCpG methylation distribution as a proxy for its “W-shape”, we observed that lymphoid cancers had significantly higher standard deviation than both peripheral blood cells and sorted B and T lymphocytes, as expected (fig 1D).

### fCpG loci are enriched in silent regions of the genome

fCpGs represented less than 1% of measured CpGs and were generally evenly dispersed across all chromosomes, with the exception of an enrichment on chromosome 19 (*P*=2.2e-5, χ^2^ test, holm-sidak (hs) correction, fig. S2A). Despite the similar distribution across chromosomes, fCpG loci were enriched on the shores of CpG islands, whilst being depleted within CpG islands themselves and the open sea (all significant p values < 10^-7^, χ^2^ test, hs correction, fig. S2B). Remarkably, fCpG were poorly overlapped with CpG previously used in a variety of “epigenetic clocks”, including mitotic age, chronological age, trait predictors and biological and mortality clocks (fig S2C, table S5, Methods).

Next, to extend our analyses beyond Illumina array data, we looked at the methylation patterns of sorted bulk B-cell and T-cell populations using publicly available whole genome bisulfite sequencing data (Loyfer et al. 2023). Methylation at fCpG loci in these polyclonal normal samples was largely intermediate (fig. S3A), consistent with our previous findings in array data (fig. 1C-D). Over a small window of 100bp, as the distance from the fCpG increased, an increasing fraction of the neighbouring CpG loci were either hyper- or hypo- methylated (fig. S3A-B), suggesting that a particular 3D DNA structure may allow the fluctuation of methylation at the specific sites where fCpGs are located.

The chromatin configuration of fCpG loci in normal and neoplastic B cells showed that fCpG loci were enriched in weak promoters and enhancers as well as H3K27me3-marked regions, whereas in active promoters and H3K36me3-marked regions they were significantly underrepresented (fig 1E). In line with this, RNA-seq analysis of 294 CLL samples showed that the expression of genes associated with fCpGs was highly skewed towards lower expression compared to genes exclusively associated with non-fCpG loci (fig 1F). Using a subset of CLL samples (n=224) having matched RNA-seq and DNA methylation data, we observed similarly low gene expression levels between fCpG-associated genes regardless of the methylation allele states (fig. 1F, Methods). These results were further supported by a variety of statistical analyses and database annotations, including a significantly lower likelihood of fCpGs being annotated as associated with a gene (fig. S2DC, p<0.001) or annotated as promoters by the ENCODE Methylation Consortium (Dunham et al. 2012) (fig. S2E p<10^-10^, χ^2^ test, hs correction). fCpG associated genes were underrepresented in pathways ubiquitously expressed across multiple tissue types (e.g. lymph node, tonsil and testis) by gene-set enrichment analysis (fig. S4A, table S6). Conversely, fCpGs were enriched in CpGs unclassified by ENCODE (p < 10^-10^, χ^2^ test, hs correction, fig. S2E) and enriched in developmental pathways (fig. S4B, sup table S7).

Together, these results indicate that fCpG loci are located in particular regions of the genome where it is likely that the DNA structure enables fluctuation, and further that fCpGs do not regulate transcription and thus are unlikely to experience clonal selection.

### EVOFLUx measures clonal evolution

We developed a stochastic modelling and inference framework called EVOFLUx to simulate how clonal evolution quantitatively determines fluctuating methylation values and enables inference of evolutionary dynamics on a case-by-case basis from patient data.

The model simulates the ongoing gain and loss of methylation at fCpGs within a lineage from a patient’s birth until the beginning of a cancer-associated clonal expansion at some specified time, and then continues to simulate methylation fluctuations within the growing population of cancer cells until the cancer sample is collected at time *T* (fig 2A; Methods and Supplementary Information). The key parameters in the model are:

- Cancer growth rate per year (*θ*), assuming an exponentially growing population.
- Patient age at cancer emergence (τ), measured in years.
- fCpG switching rates per allele per year. Four parameters corresponding to the four possible transitions between homozygous unmethylated, heterozygous methylated and homozygous methylated (*µ*, *v*, *y* and ζ).

Importantly, the time of cancer emergence should be thought of as the time of occurrence of the most recent common ancestor (MRCA) of the sampled extant cancer population, rather than the cancer age *per se*, as there may have been previous clonal expansions prior to the “final” clonal sweep that produced a clinically-detectable cancer, and the MRCA of the sample is inevitably the same or more recent than the MRCA of the whole cancer population. By combining the cancer growth rate and the time that has passed since the MRCA, the cancer effective population size (N_e_) present within the sample can be calculated as *N*_e_ *= e^θ(T- τ).^* Note that this is the *N*_e_ of the sample, rather than the cancer as a whole, and under the assumption the cancer is growing exponentially.

EVOFLUx accounts for sample impurity (contaminating non-cancer cells in the sample) by modifying the methylation fraction of the cancer cells by the average methylation of the contaminating cells (assumed to be random), weighted by sample purity (Methods). The framework also mimics the technical noise introduced by the methylation array, by adding beta-distributed noise to the methylation values centred upon the “true” methylation value in the population.

Simulations showed that the model qualitatively recapitulates the patterns observed in patient data, including the characteristic W-shape (fig. 2B). The specific shape of the resulting W-distribution is determined by the model parameters. For example, as the cancer age is increased (i.e. the time *r* occurs earlier in a patient’s life, all other parameters held constant), there is more time for fluctuations to “desynchronise” the methylation pattern of the founder cell, causing the flanking peaks of the W-shaped distribution to move towards the central peak (fig. 2C). Similarly, if we keep the time since the MRCA and the epimutation rate (per year) fixed but decrease the growth rate of the tumour population, each subsequent division has more time to accumulate epimutations, magnifying the stochasticity of the system and broadening the width of the peaks in the W-distribution (fig. 2C).

**Figure 2.**
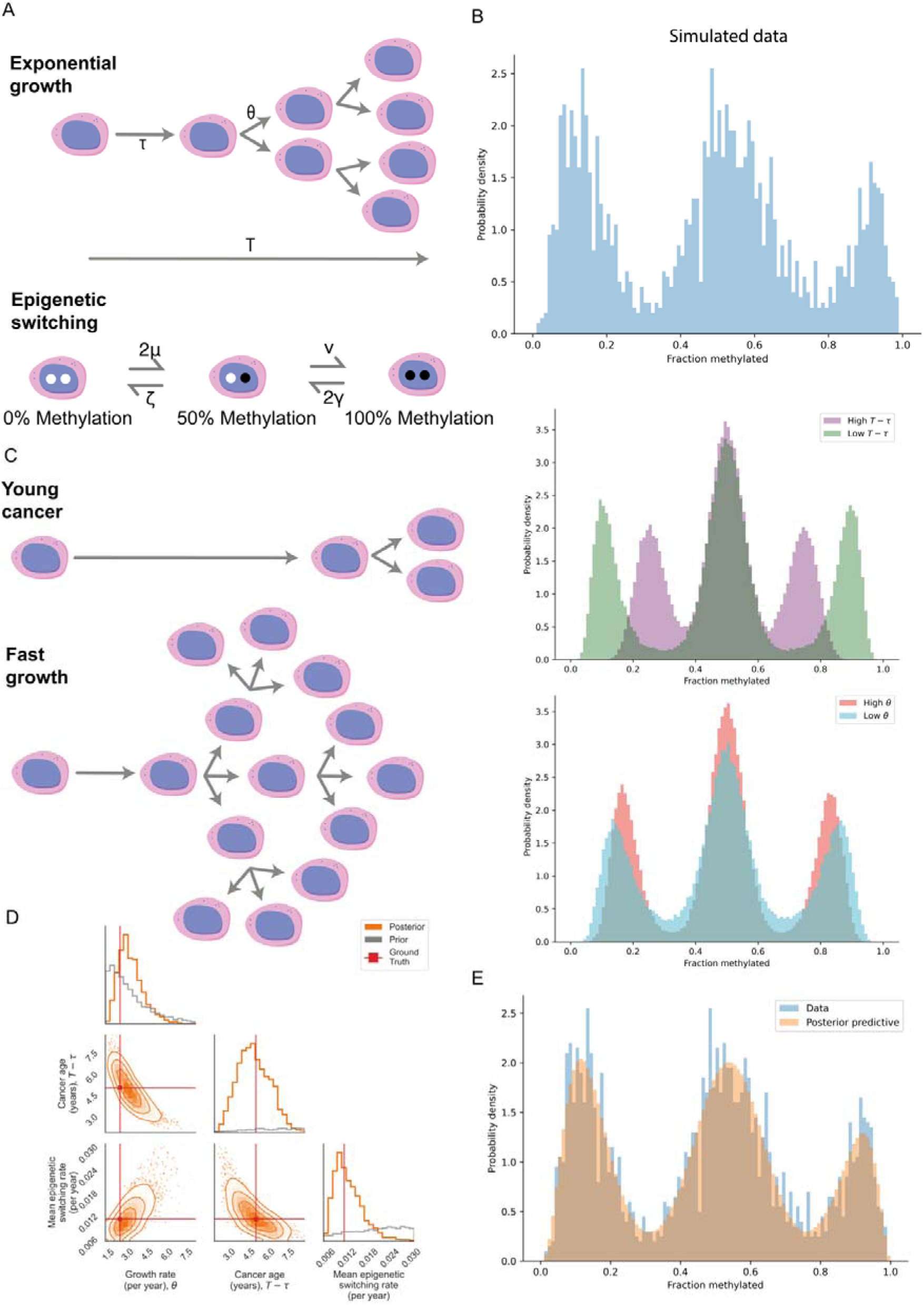
A stochastic mathematical model well-describes the observed patterns in cancer fCpG data. **A:** An illustration of the mathematical model describing how the fCpG distributions vary with the evolutionary parameters of a cancer. (top) The model is split into 2 phases, prior to the MRCA (_r_) whereby methylation changes occur in a single cell, and following the MRCA where the population grows exponentially (_0_). (bottom) At each time step, epigenetic switching is allowed to occur between the 3 possible states (rates _µ_, _v_, _(_ and _y_). **B:** An example simulation run, which has the characteristic “W-shape” observed in lymphoid cancer data (e.g. fig. 1D). **C:** Illustrative limiting cases of: (top) a very old cancer in which the fCpGs have had a long time to desynchronise vs a young cancer in which the fCpGs are largely synchronised, (bottom) a rapidly growing cancer in which the rate at which cells divide is high compared to the switching rate vs a slowly growing cancer in which the effect of stochasticity is amplified. **D:** The resulting posterior distributions after fitting the simulated data in fig. 2B with the Bayesian inference method. The posterior (orange) displays tightening around the ground truth parameter values (red) compared to the prior (grey). **E:** The posterior predictive distribution (orange) superimposed upon the simulated data (blue), showing an excellent fit.

EVOFLUx utilises an extensive Bayesian inference method to learn model parameters from input fCpG methylation distribution data (Methods). To evaluate the accuracy of the inference, we simulated fCpG data and tested the ability of EVOFLUx to recover the known input parameters (fig. 2C). These values were inferred with good confidence, with the 95% credible intervals encompassing the ground truth values (fig. 2D), significant narrowing of the posterior compared to the prior and with the posterior predictive distribution well-matching that of the original simulated dataset (fig. 2E). Hence, these simulation experiments demonstrate that the parameters defining the evolutionary dynamics of growing cancers can be measured using bulk methylation arrays.

### The evolutionary dynamics of lymphoid malignancies

We applied EVOFLUx to 1,976 samples from lymphoid cancer (including T-ALL, B-ALL, CLL, MCL, DLBCL and MM) and premalignant conditions (i.e. MBL and MGUS) where we had age and tumour cell purity information, and inferred the cancer growth rate, time to the MRCA and epigenetic switching rates of each cancer independently (table S8). Posterior distributions were well-formed (fig S5A), and posterior predictive distributions recapitulated the input data well (fig S5B), emphasising the excellent fit of the model to data.

Acute paediatric leukemias derived from precursor B/T cells and adult lymphoid neoplasms derived from mature B cells were observed to have markedly different evolutionary histories (taking the median as a point estimate; fig. 3A). In particular, EVOFLUx inferred ALLs growing at a significantly higher rate (fig. S6A, *P*= 9.3e-306, MW-U, hs correction), having a smaller cancer effective population size (*N*_e_, fig. S6B, *P*= 8.1e-25) and shorter time since the MRCA (fig. S6C, *P*= 6.0e-306) than all other lymphoid malignancies. T-ALL were found to grow faster than B-ALL (*P*=0.0017, hs correction), although the variance on the B- ALL growth rates was larger than for T-ALL (*P*=0.00044, Levene test). In adult cancers, MBL, an asymptomatic precursor condition to CLL, was found to have lower growth rates than CLL (fig. S6A, *P*= 9.7e-10) and longer time since the MRCA (*P*=9.9e-13), although there was no statistically significant difference in the inferred *N*_e_ (*P*= 0.53). DLBCL was found to have the largest *N*_e_, despite not having a particularly elevated growth rate compared to the other adult cancers (potentially due to the lower purity of DLBCL samples).

**Figure 3.**
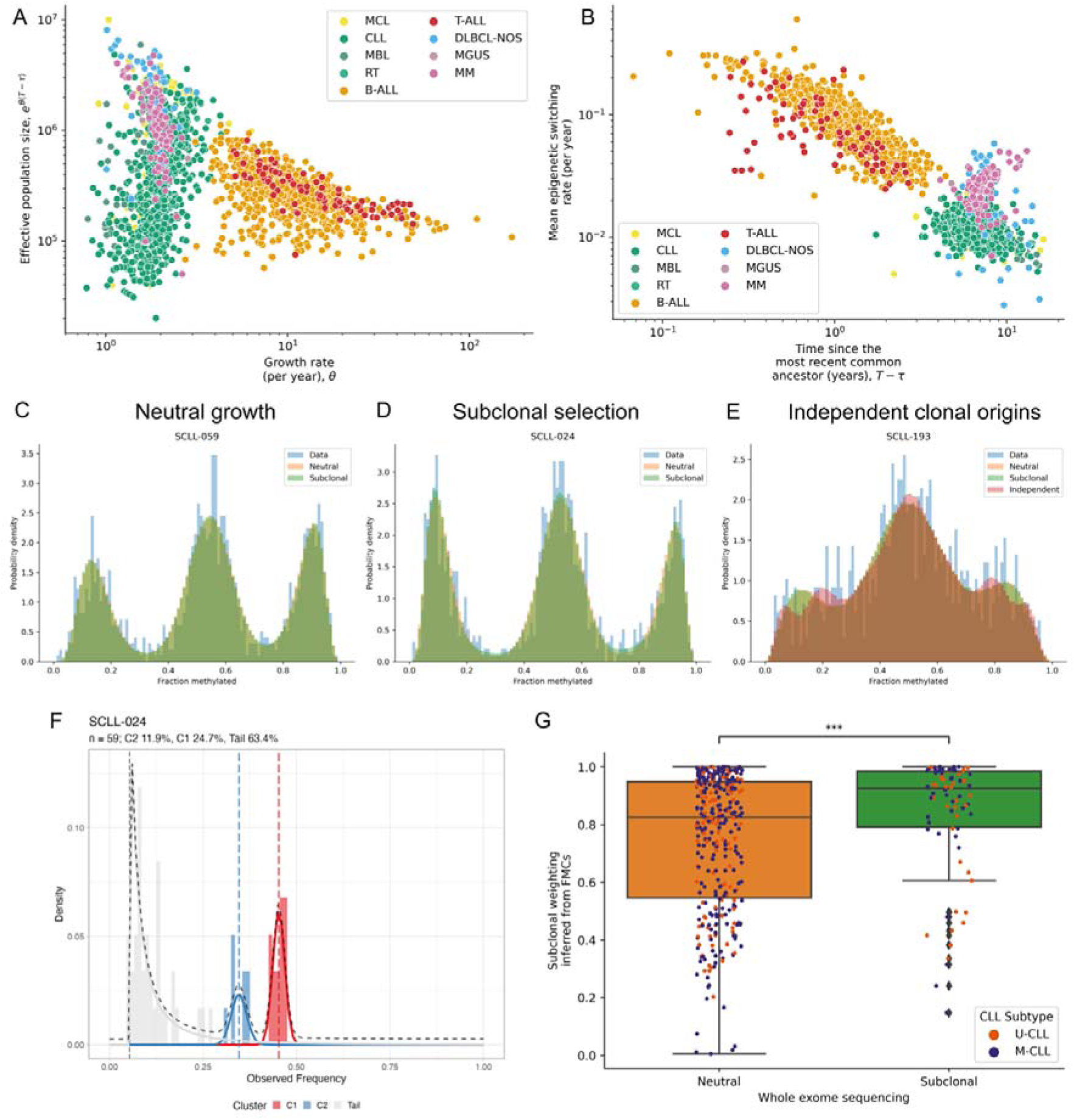
EVOFLUx reveals the evolutionary dynamics of lymphoid cancers. **A:** A scatter plot of the inferred growth rate (_0_) vs effective population size (_Ne_, _e_^B(T-r)^) per sample, using the posterior median as a descriptive summary statistic. Coloured according to cancer type. **B:** A scatter plot of the inferred time since the MRCA (_r_) vs the mean epigenetic switching rate (averaged across different rate parameters) per sample. **C-E:** 3 example histograms samples in which the neutral (C), subclonal selection (D) and independent clonal origins (E) models are preferred. **F:** A histogram showing the variant allele frequency spectrum of matched whole exome sequencing (WES) data, with the results of subclonal deconvolution (performed using Mobster (Caravagna et al. 2020)) overlaid. In this sample, there is strong evidence of an emerging subclone (C2), in line with the inference from EVOFLUx. **G:** Boxplots comparing the distribution of subclonal weightings inferred by EVOFLUx in samples called as neutral vs under subclonal selection via the WES data.

There was also significant variation in the epigenetic switching rates (epimutation rates) between diseases, with acute paediatric leukemias having an order of magnitude faster switching than adult mature B cell neoplasms (fig. 3B; fig S6D, *P*=5.6e-301). Further, we observed a positive correlation between growth rate and methylation switching rate in both B-ALL and T-ALL (fig. S6E, *P*= 2.4e-98, R^2^=0.44 & *P*=5.9e-06, R^2^=0.22 respectively), a weak negative correlation in MM (*P*=1.6e-05, R^2^=0.18) and no correlation in the other adult malignancies, suggesting that fCpG switching rates are elevated during childhood, but fall to a steady rate during adulthood. Notably, when comparing the four epigenetic switching parameters (*µ*, *v*, *y* and ζ), the different cancer types occupied different regions of the epimutation parameter space (fig. S6F).

### The Majority of CLLs Grow Neutrally

Previous work has proposed that many cancers grow effectively neutrally with no subclonal selection detectable (Sottoriva et al. 2015; M. J. Williams et al. 2016; 2018; Ling et al. 2015). In CLL, two major molecular subtypes are defined based on the extent of somatic hypermutation in the heavy-chain variable region of the IG gene, namely unmutated (U-CLL), and mutated (M-CLL). Nonetheless, a small fraction of CLL patients present with multiple IGHV rearrangements (Plevova et al. 2014), and likely consistent of two cancers with independent clonal origins. Subclonal, independent and neutral growth dynamics are distinguishable by characteristic patterns of mutations across the cancer genome (Caravagna et al. 2020; M. J. Williams et al. 2018).

Simulations showed that in fCpG data the signature of subclonal selection is the presence of intermediate peaks between the peaks corresponding to fCpG loci that were in the homozygous unmethylated, heterozygous, and homozygous methylated states in the MRCA cell (fig. S7A). These additional intermediate peaks are formed by fCpG loci that have switched methylation state in the lineage that formed the subclone within the intervening time between the MRCA of the sample population and the emergence of the outgrowing subclone. However, direct detection of these fCpG methylation patterns in the data (e.g. via a peak detection method) was generally impractical due to the relatively small number of fCpG loci that are expected to change methylation status in the time between the MRCA and the emergence of the subclone (e.g. assuming *µ* ≈ *0.01* per year, as in CLL, *1- e−2µτ2−τ1≈2%* of fCpGs in the homozygous unmethylated state will have undergone 1 fluctuation in a single year) and technical noise of fCpG methylation measured by the methylation array (fig. S7B).

Therefore, we re-fitted each of our patient’s fCpG methylation distributions with modified EVOFLUx models that included subclonal selection and independent clonal origins, and used Bayesian model selection and a leave-one-out approach to determine which model best represented the data (fig. 3C-E, S7C, Methods). We found that in the majority of samples (1610/1976), there was no evidence of either subclonal or two independent cancers at 95% confidence. There was also a high variance in the fraction of cancers growing under subclonal selection between different cancer types (fig. S8A, from over 30% (232/718) in CLL to less than 5% in DLBCL (1/57). However, these results need to be interpreted cautiously given the heterogeneity in purity across the sample groups, which likely limits the power to detect subclonal selection in lower purity samples.

We note that only strong selection occurring neither “too early” or “too late” is detectable in the data (fig. S7D), analogously to detecting selection using point mutations in WGS (Heide et al. 2018). This is because late arising or weakly selected subclones will only be present at very low frequencies, whereas very strongly selected early subclones will tend to have swept prior to sampling, and very early arising subclones have not had sufficient time to evolve fCpG patterns dissimilar enough to be distinguishable from the ancestor clone. Furthermore, whilst our simulations demonstrated that EVOFLUx can correctly identify subclonal section, the absolute value of the inferred selection coefficients are not well informed due to the noisiness of the methylation array data (fig. S7E).

To verify EVOFLUx based inferences of subclonal selection we used matched WES data available for 425 CLL cases. Using MOBSTER, our subclonal deconvolution tool (Caravagna et al. 2020), to detect emerging subclone(s) from the variant allele frequency (VAF) distributions of somatic mutations (fig. 3F, table S9) we found 78 /425 samples contained evidence of subclonality by WES (table S10). Samples which were classified as subclonal by MOBSTER had significantly higher subclonality weights inferred from our EVOFLUx inference (*P*= 2.0e-4, MW-U test, fig. 3G).

The paucity of point mutations in WES data limits the power to detect subclonality. MOBSTER was more likely to assign a sample as containing a subclone when the sample contained more mutations (p= 0.0023, logistic regression against log number of mutations, fig. S8B). Hence, we suggest that those samples with high subclonality weights from our EVOFLUx inference, despite being labelled as “neutral” by MOBSTER, may truly contain subclones. Conversely, in cases assigned as subclonal by MOBSTER, there was no association between the mutation burden and EVOFLUx subclonality weighting, suggesting these may be genuinely subclonal samples that EVOFLUx has missed (*P*=0.24, logistic regression, fig. S8C).

Cancers with two independent clonal origins were detected in 22/718 CLL cases. We attempted to validate these inferences by comparing them to orthogonal measurement of the number of IG gene rearrangements based on WES/WGS and RNA-seq (Knisbacher 2022) (table S11). Those patients with multiple IG gene rearrangements had elevated independent origins model weightings compared to those without (fig S8D, *P*= 0.028), although we note that EVOFLUx was not as accurate at detecting independent cancers as detecting subclonality.

#### EVOFLUx detects early subclonal origins of Richter transformed cells in CLL

A small fraction of CLL patients will undergo RT, the emergence of an aggressive phenotype with dismal prognosis. Recently, using single cell genomics and transcriptomics, we found that RT cells are present at low frequencies within the diagnostic CLL sample, preceding the clinical emergence of RT in some cases by two decades (Nadeu et al. 2022). For 2 of the same patients analysed previously by WGS and single cell approaches (Nadeu et al. 2022), we also had DNA methylation arrays of longitudinal samples from diagnosis to RT (fig 4A, table S12). We used EVOFLUX to construct phylogenetic relationships between samples from differences in fCpG methylation (Methods), extracting valuable temporal information regarding the divergence of RT cells from the ancestral clone.

We found the inferred phylogeny well matched the known clinical relationships between the different samples (fig. 4B). Notably, the evolutionary distance between two technical replicates of untreated samples (450k and EPIC data) was small in both cases (fig. 4B). In both phylogenies, the RT clone diverged exceptionally early, roughly a decade before the MRCA of the non-transformed samples (9 and 12 years for case 12 and 19, respectively). This is consistent with our previous work (Nadeu et al. 2022) in which we detected RT cells within the diagnostic sample, but suggests that the initial RT divergence occurred well before diagnosis, over 30 years before the clinical presentation of RT.

**Figure 4.**
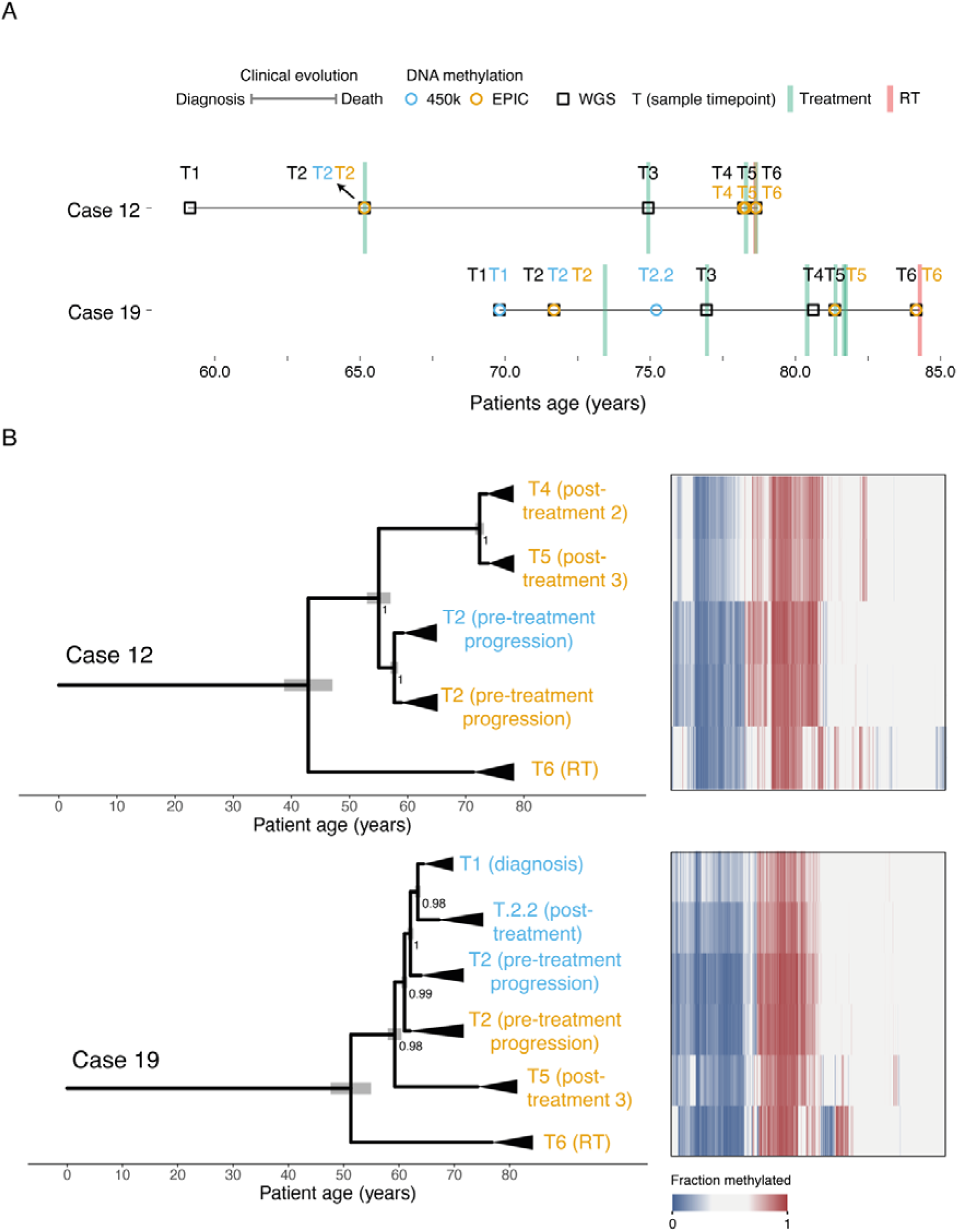
fCpGs allow for phylogenetic reconstruction of longitudinal lymphoid cancer samples. **A:** Timeline of two CLL patients with samples collected longitudinally, annotated with treatment received. Circles represent methylation array (blue: 450K, orange EPIC), squares represent whole genome sequencing, treatment is represented with a green vertical line and Richter transformation (RT) is represented with a vertical pink line. **B:** (left) The reconstructed phylogenies of the relationship between samples, annotated with the clinical classification of each sample. The black triangles represent the time that occurred since the most recent common ancestor, taken as the posterior median of _T-r_ from the single-sample EVOFLUx inferences. (right) A heatmap representing the 978 fCpG loci, with the colour a representing the fraction methylated (0% blue, 100% red).

To validate the fCpG phylogenetic inference, we built phylogenetic trees on SNV data from matched WGS (Nadeu et al. 2022) (fig. 4A and S9, table S12). We restricted our analysis to clock-like SBS1 mutations (Nik-Zainal et al. 2012) which were clonal within the sample (Caravagna et al. 2020). The topology of the SNV phylogenies matched that of the fCpG phylogenies and the 95% credible intervals of the estimated MRCA overlapped. Hence, fCpGs have the potential to serve as a high temporal resolution phylogenetic character, enabling multi-sample phylogenetic reconstruction at a fraction of the cost of WGS.

In addition, we applied our phylogenetic analysis to 3 longitudinal B-ALL samples, tracking the disease course from initial diagnosis to relapse using the same methodology (fig. S10). In each of the patients, the subsequent relapse samples formed a separate clade from the initial diagnostic samples, suggesting the cancer population has undergone a significant bottleneck through treatment. The branch lengths of the diagnostic samples were all very short, suggesting there was initially little evolutionary distance between the surviving resistant population and the sensitive cells.

#### Evolutionary growth dynamics of CLL predicts clinical outcomes

Since cancer development and progression is fundamentally an evolutionary process, the evolutionary trajectory of the disease should predict the future course of the disease and clinical outcome of the patients. To evaluate this hypothesis, we first compared the inferred growth rate and *N*_e_ between different molecular subtypes of the same cancer type.

In B-ALL, cases having the translocation 11q23/MLL had a significantly higher growth rate (fig. 5A, *P*=1.3e-13, MW-U, 44.3±6.1 vs 11.7±0.2 per year [mean ± standard error]), but lower *N*_e_ (*P*=3.7e-07, 1.8e5±0.2e5 vs 3.0e5±0.06e5 cells) than the other subtypes, consistent with its distinct clinical behaviour (Rice and Roy 2020). In MCL, the generally more indolent nnMCL (Nadeu et al. 2020) had a lower growth rate (fig. 5B, *P*=1.1e-03, 1.7±0.1 vs 2.1±0.1 per year) and N_e_ (*P*=7.4e-05, 4.7e5±1.4e5 vs 1.5e6±0.2e6 cells) than the more aggressive conventional MCL (cMCL). In DLBCL transcriptomic subtypes (Alizadeh et al. 2000) there was no significant differences, likely due to the smaller number of cases and the lower sample purity (fig. S11A&B).

**Figure 5.**
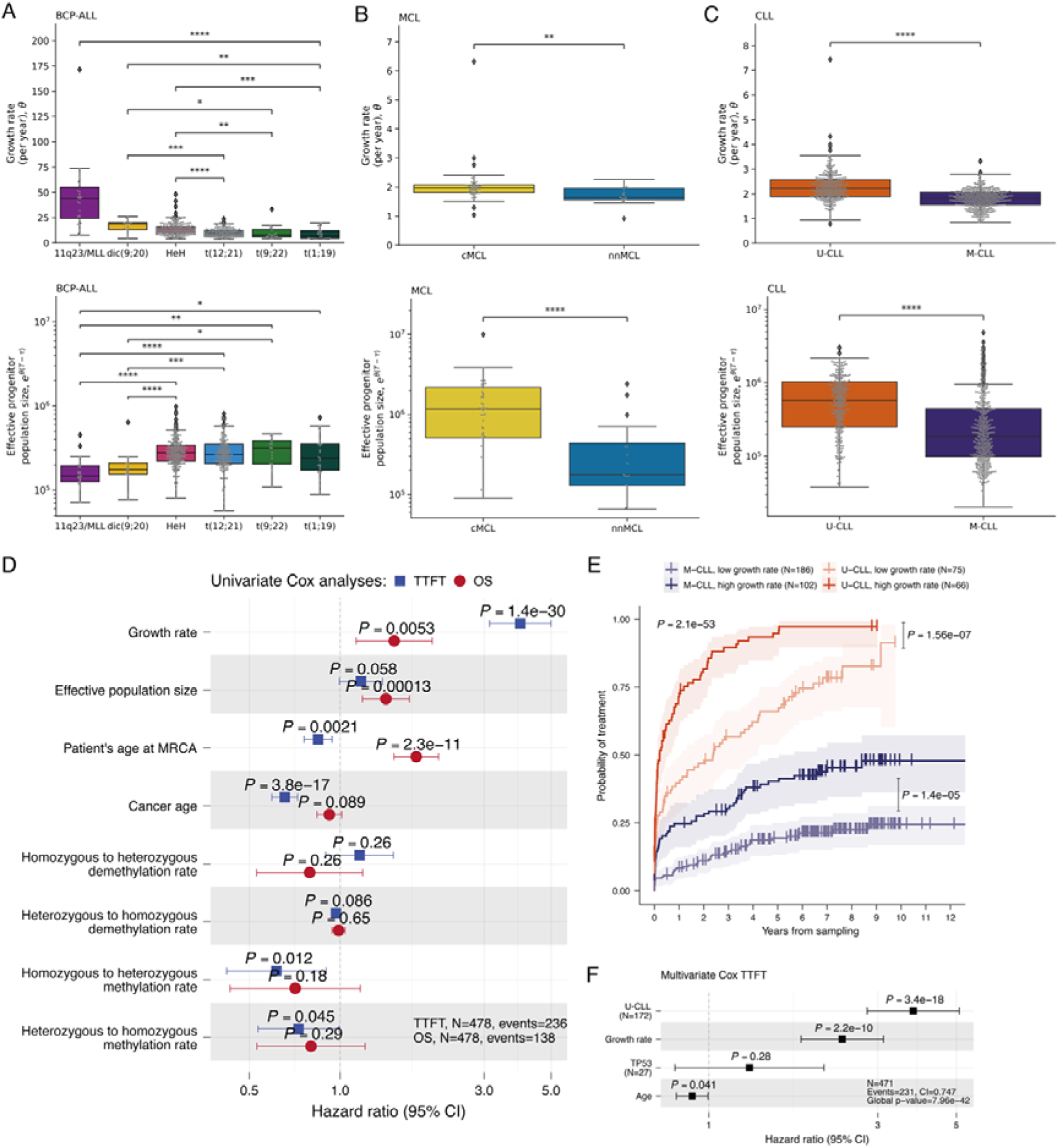
The evolutionary history of a tumour varies between molecular subtype and is predictive of clinical outcome. **A-C:** The inferred growth rate (top) and effective population size (N_e_, bottom) of individual cancer samples separated by molecular subtype in B-acute lymphoblastic leukaemia (A, BCP-ALL), mantle cell lymphoma (B, MCL) and chronic lymphocytic leukaemia (C, CLL). Differences between subtypes were tested using Mann-Whitney U tests, with Holm- Bonferroni corrections applied. In A, the 11q23/MLL subtype had significantly higher growth rate than each of the other B-ALL subtypes (all _P_<0.005), but for ease of presentation, only the comparison with t(1;19) is shown here. **D:** Univariate survival analysis of the time to first treatment (TTFT, blue) and overall survival (OS, red) in the discovery CLL cohort for evolutionary variables inferred via EVOFLUx. **E:** Kaplan-Meier curves comparing the TTFT between patients with high vs low inferred cancer growth rates, separated by IGHV mutational status. **F:** Multivariate Cox regression of the TTFT shows the cancer growth rate is significant when controlling for IGHV status, TP53 status and age.

In CLL, the more aggressive U-CLL subtype (Hamblin et al. 1999; Damle et al. 1999) showed a significantly higher growth rates (fig. 5C, *P*=1.3e-32, 2.3±0.04 vs 1.8±0.02 per year) and *N*_e_ (*P*=2.1e-22, 7.2e5±0.3e5 vs 4.1e5±0.3e5 cells) compared to M-CLL. In its precursor condition MBL, although we had little power to detect a difference between U-CLL and M-CLL subtypes, we could observe a non-significant increase in growth rate in U-CLL cases mirroring the findings in CLL (fig. S11C&D, *P*=0.093, 1.7±0.1 vs 1.5±0.06 per year). This result is consistent with the known enrichment of MBLs in IGHV mutated cases, and suggestive that MBL patients with an unmutated IGHV gene generally progress more rapidly towards CLL and therefore the MBL phase is less frequently observed in the clinical setting.

To further measure the influence the past evolutionary history of a cancer imparts upon its future clinical course, we exploited our access to a large clinically well-annotated series of CLL cases to evaluate the prognostic value of the inferred evolutionary parameters. We used univariate Cox models to assess the impact of each parameter on time to first treatment (TTFT), a commonly used variable that describes the natural history (biological behaviour) of the CLL cells, as well as overall survival (OS) which is a more complex endpoint as it convolves disease biology with heterogeneous treatments administered to the patients.

The univariate prognostic impact of the inferred growth rate of the cancer on both TTFT (fig. 5D, *P*=1.4e-30, hazard ratio (HR) HR=3.95) and OS (*P*=0.0053, HR=1.51) was stark. The *N*_e_ of a cancer did not have a strong impact on TTFT (*P*=0.058, HR=1.17), but its effect on OS was clear (*P*=1.3e-4, HR=1.41). The patient age at the time of the MRCA of the CLL population is highly corelated with the age of the patient (fig S12A), so unsurprisingly older patients had worse OS (*P*=2.3e-11, HR=1.79). The decrease in risk of progression with cancer age (*P*=3.8e- 17, HR=0.65), measured by the time since the MRCA, was likely due to confounding with the growth rate, as these parameters were negatively correlated (fig. S12A). The epigenetic switching rate parameters were largely uninformative of prognosis.

As the growth rate was different between U-CLL and M-CLL (fig. 5C), we next analysed its prognostic impact within each of them. Indeed, we observed that the estimated growth rate has a profound impact on TTFT in both CLL subtypes separately (fig. 5E, *P*=1.4e-5 for M-CLL, *P*=1.56e-7 for U-CLL, overall *P*=2.1e-53). Cases with higher growth rate consistently showed a shorter TTFT. Considering the growth rate as continuous variable in a multivariate Cox regression model, it maintained a strong independent prognostic impact (*P*=2.2e-10, HR=2.28) in the context of other well-established clinical variables, including the IGHV mutational status and *TP53* aberrations (including both mutations and deletions) and age at sampling (fig. 5F). The cancer *N*_e_ was more significantly correlated with OS than the growth rate, and this effect was preserved in the U-CLL subtype (fig S12B, *P*=0.55 for M-CLL, *P*=1.0e- 6 for U-CLL, overall *P*=1.7e-9), and in multivariate setting with IGHV status, *TP53* aberrations and age at sampling (fig S12C, *P*=0.025, HR=1.33).

The inference of the evolutionary parameters on our initial cohort was wholly blinded to the clinical outcomes. Nevertheless, we validated the prognostic value of the inferred evolutionary parameters using a second independent cohort of 210 CLL patients (136 untreated at sampling; table S2) (Oakes et al. 2016; Dietrich et al. 2018). We found that the growth rate was prognostically relevant as a continuous (fig. S13A, *P*=7.4e-4, HR=2.39) and dichotomic variable (fig. S13B, *P*=5.1e-5). In a multivariate Cox model with IGHV, *TP53* status and age the growth rate approached significance (fig. S13C, *P*=0.11, HR=1.62). We note that the power of the study in this cohort was limited, as even the well-validated IGHV status had borderline significance (*P*=0.053). In this cohort, the predictive impact of *N*_e_ on OS was not found to be significant (fig. S13A, *P*=0.25, HR=1.39), but we stress that TTFT is a better reflection of the natural history of disease, not confounded by heterogenous treatment decisions.

These results demonstrate that the evolutionary parameters inferred from a cost-effective methylation array could have a direct clinical application by contributing to predict the clinical behaviour of CLL patients independently from well-established prognostic variables.

## Discussion

Our study establishes a framework, called EVOFLUx, that enables quantitative measurement of the evolutionary dynamics of human malignancies. These dynamics represent the long-term evolutionary history of the neoplasm and are fundamentally distinct from characterisations of the contemporary phenotype of cancer cells, such as the fraction of proliferating cells. The framework only requires data obtained from single-timepoint bulk tumour samples that have been processed with widely-available and low-cost DNA methylation arrays as input. The methodology should also work identically for sequencing-based measurement of DNA methylation, should generalise across cancer types, and be applicable to proxy tumour measures such as tumour-derived cell free DNA (cfDNA) extracted from blood. Consequently, this strategy enables measurement of evolutionary dynamics at massive scale, which paves the way towards clinical translation.

EVOFLUx utilises the clonal lineage identity signal that derives from stochastic fluctuations in methylated and unmethylated states at a subset of CpG sites across the genome, and outputs the evolutionary parameters defining the longitudinal dynamics of cancer growth, alongside precise measurement of DNA methylation and demethylation rates at these sites. Epimutation rates were found to differ by more than an order of magnitude between paediatric and adult malignancies. Thus, we suggest that future analyses of these quantitative data are likely to reveal new mechanistic insight into the somatic maintenance of DNA methylation.

Applying our framework in lymphoid malignancies reveals that cancer evolutionary dynamics are strongly associated with patient clinical outcomes across disease types, and further that evolutionary dynamics add significant new prognostic value to state-of-the-art approaches currently applied to the management of CLL in the clinical setting. We consider this strong evidence that clonal evolution, the fundamental cellular process of disease development, underlies the clinical course of the disease. Further, as cancer development and progression are in essence evolutionary processes, we expect these results to generalise across all cancer types. We note that genome-wide DNA methylation analyses also measure other important biological features of a cancer (for example in CLL: (Duran-Ferrer et al. 2020)), that could be combined with EVOFLUx-based inference of evolutionary dynamics to further improve the prognostic value of DNA methylation data.

In summary, we present a cost-effective high-throughput platform for measuring cancer evolutionary dynamics in patient samples. These fundamental measurements of the disease biology hold significant prognostic value and represent an innovative asset in the field of precision oncology.

## Author Contributions

TAG and JIMS conceived, funded, and supervised the study. CG and TAG conceived and designed the modelling scheme and the Bayesian inference framework, with support from CPB, and CG implemented and ran this. CG, MDF, HG, JH and FN performed bioinformatic analysis. MDF and JIMS performed survival analysis. CG, HG and DM designed and ran the phylogenetic analysis. MDF, FN, NV, OK, JN, TZ, EC and ALG provided data and contributed to sample biological and/or clinical annotation. MDF, NV, JF, DS and JIMS provided clinical insight. Our study builds upon DS’s earlier conception of the fluctuating methylation phenomenon. CG, MDF, JIMS and TAG wrote the manuscript, and all authors approved the final version.

## Data Availability

No new data was generated in the course of this study. The harmonised and filtered methylation matrix is available (preferred browser Firefox) at: http://inb-cg.bsc.es/hcli_priv/publications_data/fluctuating_cpgs_gabbutt_duran_ferrer_2023/QC_2204_ssNob.tsv.7z - the password for this file is available upon request to the authors.
Previously published DNA methylation data re-analysed in this study can be found under accession codes: B cells, EGAS00001001196; ALL, GSE56602, GSE49032, GSE76585, GSE69229; MCL, EGAS00001001637, EGAS00001004165; CLL, EGAD00010000871, EGAD00010000948, EGAD00010001975; MM, EGAS00001000841; DLBCL, EGAD00010001974.
CLL gene expression data is available EGAS00001000374 and EGAS00001001306. ChIP-seq datasets are available from Blueprint https://www.blueprint-epigenome.eu/ under the accession EGAS00001000326.
Matched WES and WGS are available under accessions EGAS00000000092 and EGAD00001008954 respectively.

https://ega-archive.org/studies/EGAS00001001196

https://www.ncbi.nlm.nih.gov/geo/query/acc.cgi?acc=GSE56602

https://www.ncbi.nlm.nih.gov/geo/query/acc.cgi?acc=GSE49032

https://www.ncbi.nlm.nih.gov/geo/query/acc.cgi?acc=GSE76585

https://www.ncbi.nlm.nih.gov/geo/query/acc.cgi?acc=GSE69229

https://ega-archive.org/studies/EGAS00001001637

https://ega-archive.org/studies/EGAS00001004165

https://ega-archive.org/datasets/EGAD00010000871

https://ega-archive.org/datasets/EGAD00010000948

## Acknowledgements

This research was primarily funded by an Accelerator award Cancer Research UK/AIRC/AECC joint funder-partnership (to JF, JIMS and TAG) and the US National Institutes of Health National Cancer Institute (U54 CA217376, to DS and TAG). Additional funding: Cancer Research UK (A19771 and DRCNPG-May21_100001) to TAG, the European Research Council under the European Union’s Horizon 2020 Research and Innovation Program (810287, BCLLatlas, to EC and JIMS), La Caixa Foundation (CLLEvolution LCF/PR/HR17/52150017 [HR17- 00221LCF] and CLLSYSTEMS - LCF/PR/HR22/52420015 [HR22-00172] Health Research 2017 and 2022 Programs, to EC), Generalitat de Catalunya Suport Grups de Recerca AGAUR (2021-SGR-01343 to JIMS and 2021-SGR-01172 to EC). JN and OK were supported by the Swedish Research Council (2019-01976), Swedish Childhood Cancer Fund (PR2019-0046/PR2022-0082), and Swedish Cancer Society (CAN2022-2395). CG was further supported by the BBSRC London Interdisciplinary Doctoral Programme (LIDo) and Thorton Foundation funding to the Institute of Cancer Research. MDF was supported by a postdoctoral fellowship of the AECC Scientific Foundation. This research utilized the Cancer Research UK City of London High Performance Computing (HPC) facility, and was partially developed at the Centro Esther Koplowitz (CEK, Barcelona, Spain). We acknowledge the SciLifeLab National Genomics Infrastructure, SNP&SEQ Technology Platform in Upppsala, Sweden, funded by the Swedish Research Council and the Knut and Alice Wallenberg Foundation, for assistance with DNA methylation analyses.

## Methods

### Assembly and quality control of DNA methylation data

We assembled and processed with a harmonized pipeline (Duran-Ferrer et al. 2020) (version 4.1, see code availability section) 2,430 bulk sample Illumina methylation array data of normal and neoplastic lymphoid cells from previous publications (Kulis et al. 2015; Nordlund et al. 2013; Reinius et al. 2012; Lee et al. 2015; Queirós et al. 2016; Nadeu et al. 2020; Duran-Ferrer et al. 2020; Nadeu et al. 2022; Oakes et al. 2016; Dietrich et al. 2018; Agirre et al. 2015). Briefly, raw idat files were loaded and processed with R (version 4.3.1) using *minfi* package (Aryee et al. 2014; Fortin, Triche Jr, and Hansen 2017) (version 1.46.0) in batches as specified in the column “SSNOB_NORMALIZATION_BATCH” of table S2. Briefly, the data was processed for each batch as follows. First, idats files were loaded into a *RGChannelSet* object, and *minfi* quality metrics using the *qcReport* function were performed, controlling for biased distributions of methylation values and low signal intensities of internal control probes for each sample. Next, further quality metrics were derived using the function *minfiQC* on unnormalized *RGChannelSet* obejct. Those samples with median signal intensities of unmethylated and methylated channels of at least 10.5 in log2 scale were considered as having good signal intensities (default value in *minfi*). Subsequently, detection *P*-values were calculated across all CpGs and samples using the *detectionP* function for the unnormalized *RGChannelSet* object. Samples were considered as good if having a mean detection P-value across all CpGs pf *P*≤0.01. On a CpG level, we retained CpGs with a detection *P*-value ≤1e^-16^ in ≥90% of the samples. The *RGChannelSet* object was normalized with the *preprocessNoob* function. We next retained only CpGs (excluding CH probes) that did not contain any single nucleotide polymorphism neither in the interrogated CpGs nor in the probe using the *dropMethylationLoci* and *dropLociWithSnps* functions. Furthermore, CpGs with any previous evidence of potential cross-hybridization were excluded (Chen et al. 2013) and only CpGs mapping to autosomal chromosomes were subsequently retained for downstream analyses. Finally, we checked the distribution of normalized methylation values and performed principal component analyses separately for samples passing all quality checks as well as those considered to as bad samples. The final DNA methylation matrix contained 2,204 samples and 389,180 CpGs passing all the aforementioned quality controls, and included 2,054 patients (22 technical replicates, 3 synchronic and 125 longitudinal samples from the same patients).

To determine to purity of samples, we used our previously deconvolution strategy to infer tumor cell content by DNA methylation (Duran-Ferrer et al. 2020), which was used as a consensus purity in all the tumor samples except for DLBCL and MM. In these two tumor entities, we have previously identified a DNA methylation signature loss causing inaccurate tumor purity predictions using DNA methylation data, and therefore we used available genetic or flow cytometry data for DLBCL and MM, respectively.

### Statistical analysis

Statistical tests performed throughout the study were performed as two-sided. Appropriate multiple test correction, such as the holm-sidak (hs) correction, is noted when applied.

### Characterisation of fCpGs

To characterise the genomic and regulatory context of fCpGs we employed a series of statistical analyses and database annotations. We annotated fCpGs using Illumina manifest and other genomic annotationa available at Bioconductor packages *IlluminaHumanMethylation450kanno.ilmn12.hg19* (version 0.6.1) and *IlluminaHumanMethylationEPICanno.ilm10b2.hg19* (version 0.6.0). We used Chi-squared tests *x*^2^ to assess the enrichment of fCpGs in distinct genomic regions or elements. We also used CpG annotations of the regulatory features provided by the ENCODE Methylation Consortium (Dunham et al. 2012). We performed gene-set enrichment analysis upon the fCpG-associated genes using gProfiler (Raudvere et al. 2019), specifically focusing upon the Gene Ontology biological processes (Ashburner et al. 2000) and the Human Protein Atlas (Uhlén et al. 2015). The statistical domain space was limited to genes targeted by at least one CpG in the 389,180 candidate CpG set and significance was determined using the g:SCS algorithm (Reimand et al. 2007). Previous chromatin segmentation of normal and neoplastic B cells was used to assess the chromatin state enrichment of fCpG (Duran-Ferrer et al. 2020; Beekman et al. 2018).

fCpG were checked for their overlap with previous “epigenetic clocks”, including mitotic (Duran-Ferrer et al. 2020; Yang et al. 2016; Teschendorff 2020; Youn and Wang 2018; Zhou et al. 2018), chronological age (Bocklandt et al. 2011; Garagnani et al. 2012; Hannum et al. 2013; Horvath 2013; Lin et al. 2016; Vidal-Bralo, Lopez-Golan, and Gonzalez 2016; Weidner et al. 2014; Zhang et al. 2019; Horvath et al. 2018; Shireby et al. 2020), gestational age (Bohlin et al. 2016; Knight et al. 2016; Y. Lee et al. 2019; Mayne et al. 2017; McEwen et al. 2020), biological age and mortality (Belsky et al. 2022; Levine et al. 2018; Lu et al. 2019), and trait predictors (McCartney et al. 2018; Liang et al. 2020). The package *methylCIPHER* (https://github.com/MorganLevineLab/methylCIPHER) was used to integrate the majority of the epigenetic clocks.

### CLL RNA-seq data

Previously available RNA-seq data for 294 CLL patients was obtained (Puente et al. 2015) and processed as previously described (Nadeu et al. 2020). Matched RNA-seq data and DNA methylation data for the same patients at the same timepoint was available for 224 CLL patients. Transcript per million counts (tpms) were used to represent differential gene expression values across genes and samples. We used the gene annotation provided in the R Bioconductor package *IlluminaHumanMethylationEPICanno.ilm10b2.hg19* to classify genes associated with fCpGs. Genes targeted by any fCpG were considered as “fCpG genes”.

In each methylation sample, the 978 fCpG were discretised as homozygous demethylated, heterozygous methylated or homozygous methylated (coded as [0,1,2] respectively). This was done by separately fitting a beta mixture model with 3 components to each sample using Stan (Carpenter et al. 2017) and extracting the component mixture probability. The gene expression value for genes classified as having and fCpG with 0, 1 or 2 alleles methylated were plotted as previously described.

### A Stochastic Model of how fCpGs vary in large, neutrally growing populations of cells

We built a generative computational model of how the patterns of fCpGs vary over time (*t*) according to the evolutionary history of a cancer. Initially, our model focused on neutral evolution, before expanding to non-neutral modes of tumour evolution below. For the full explanation of the model, please see the supplementary information.

Our model was parameterised in terms of the age of the patient at which the most recent common ancestor (MRCA) emerged (*τ*), the exponential growth rate of the cancer (*θ*), and the epigenetic switching rates of the fCpGs (*µ*, *y*, *v*, ζ). The model was partitioned into 2 phases, prior and post the emergence of the MRCA. At time *t = 0*, the fCpGs were assumed to be equally likely to be homozygously methylated or demethylated. The fCpG status of the MRCA at time *t = τ* was calculated by applying matrix exponentiation.

The second phase of the model consisted of a discrete time Markov process. The effective population size of the growing cancer was modelled as growing according to a deterministic exponential growth equation, *N*_e_ *= e^θ(t-τ).^* Each fCpG was considered independently – at each time step, *t ➔ t + δt*, the number of homozygous methylated (*m*), heterozygous methylated (*k*) and homozygous demethylated cells (*w*) at a specific fCpG was updated according to the epigenetic switching rates.

At the time of sample, *T*, the fraction methylation of each simulated fCpG was calculated by summing the number of methylated alleles and normalising by the total number of alleles in the population:

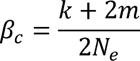

We further accounted for contaminating normal cells and the technical noise introduced by the methylation bead array. The methylation of the contaminated samples was assumed to be an average of the cancer methylation, *β*_c_*(t)*, weighted by the tumour purity *p*, and the average of the normal population, *β_n_*, weighted by *1- p*. Following our previous work, the bead array was assumed to saturate at extreme methylation values, shifting the minimum and maximum methylation by *δ* and *ε* respectively. The noise of the bead array was assumed to be beta distributed, with precision parameter *K*.

### Non-neutral models of tumour evolution

Alongside our model of neutral exponentially growing cancer populations, we devised two alternative models of cancer growth:

1. A subclonal selection model in which a single cell within the cancer develops a selective advantage and begins to grow at an increased growth rate.
2. An independent clonal origins model, whereby a patient has developed two distinct cancers concurrently.

For the subclonal selection model, we replaced the growth rate *(θ)* and the time of the MRCA (*τ*) with the growth rates and time of the MRCA of the initial, slower-growing population (*θ*_1_ and *τ*_1_ respectively), and that of the more recently emerging, faster-growing population (*θ*_2_ and *τ*_2_), constraining *τ*_1_ *< τ*_2_ and *θ*_1_ *< θ*_2_ (fig. S7C). We assumed that the initial cancer population begins exponentially growing at *r*_1_ as above, but at time *t = τ*_2_ we select a single cell with a set of fCpG states drawn according to the cancer population and allow this second population to grow concurrently with a growth rate *θ*_2_.

The independent-cancer model followed the same scheme as the nested subclonal selection model, except the methylation status of the emerging cancer was that of an independent cell which experienced random fluctuations between *t = 0* and *t = τ*_2_.

If we let the number of cells in the less fit subclone in each methylation state be *{m*_1_*, k*_1_*, w*_1_*}* and in the fitter subclone be *{m*_2_*, k*_2_*, w*_2_ *}*, following the convention above, then in both cases the measured methylation patterns at the time of sample are:

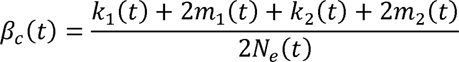

Where 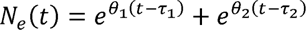.

### Prior functions

For each methylation array blood sample, we had matched age (*T*) and purity (*p*) information. Hence, the parameters to be inferred are the growth rate (*θ*), the age of the patient when the MRCA emerged (τ), the epigenetic switching rates (*µ*, *v*, *y*, ζ; defined in fig 2A), the average fraction methylated of contaminating normal cells (*β*_n_), the beta offsets from 0 and 1 due to the background noise on the methylation array (*Δ* and ɛ, respectively), and the precision of the beta distributed noise (ϰ).

These parameters are constrained either to be positive (*θ, µ, v, y,* ζ, ϰ *> 0*) or to lie within a specified range 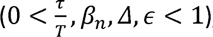, which we achieved using appropriate prior distributions. To better allow for priors to be set on a biologically meaningful scale, the priors for the lognormal distribution were set in terms of the real scale mean and standard deviation, rather than the standard log-scale. To reduce correlations in the posterior and make sampling more efficient, the variables *v* and ζ were normalised by *µ* and *y* respectively.

The priors are as follows:

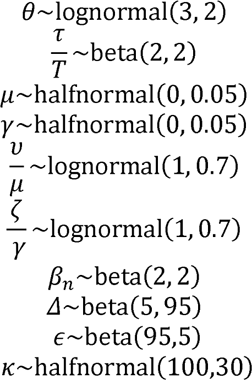

When fitting non-neutral models of tumour growth, the inference is parameterised in terms of the relative growth of the fitter subclone, 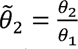, and the fraction of the population consisting of the fitter subclone, 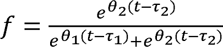. The age at which the second clone emerges is then:

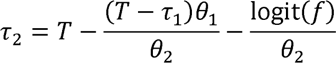

This parameterisation induces less correlations in the resulting posterior, which significantly improves the sampling efficiency. The priors on these additional parameters are:

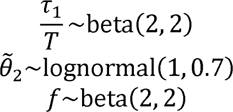

All the other priors were the same as in the neutral case.

### Bayesian Inference

We developed a stochastic estimator of the loglikelihood function at a given set of parameters by simulating the fCpG methylation distribution a large number of times, correcting for the bias inherent with using a finite number of simulations and penalising the loglikelihood for extreme values of the N_e_ (see supplementary for details).

The standard Bayesian algorithms developed to infer the posterior for a given set of data (e.g. MCMC, nested sampling) are typically used when the log-likelihood is analytically tractable and can be calculated exactly. Remarkably, it has been shown that, as long as the stochastic approximation of the log-likelihood is unbiased, MCMC methods can obtain an exact Bayesian inference of the true posterior, as in pseudo-marginal Metropolis–Hastings (Andrieu and Roberts 2009).

Here, we employed a nested sampling approach using the dynesty package (Skilling 2004; 2006; Speagle 2019). Unlike pseudo-marginal Metropolis–Hastings, nested sampling is able to efficiently explore multi-modal posterior landscapes (which can occur under the subclonal and independent cancer models).

### Model comparison between non-neutral models of tumour evolution

We employed an expected log pointwise predictive density (ELPD) (Vehtari, Gelman, and Gabry 2015) approach to compare our competing models of evolution for each sample using the arviz Python package (Kumar et al. 2019), which uses PSIS-LOO-CV to compare the out-of-sample prediction accuracy between models whilst naturally penalising more complex models. This required the loglikelihood per datapoint and the posterior predictive for every point in the posterior. The weights of the respective models were calculated using pseudo-Bayesian Model averaging using Akaike-type weighting, stabilized using the Bayesian bootstrap (Yao et al. 2017).

### CLL and RT genomic analyses

Previous mutated annotation files (MAF) from WES data (Knisbacher et al. 2022) and WGS (Nadeu et al. 2022) were used to further validate our distinct EVOFLUx evolutionary modes (i.e., neutral, subclonal and independent) and RT phylogenies.

### Subclonal deconvolution of WES and WGS data

To detect subclones in bulk WES and WGS data, we employed MOBSTER (Caravagna et al. 2020), which fits the variant allele frequency (VAF) spectrum with a mixture model containing a Pareto distribution to account for the neutral tail (M. J. Williams et al. 2016) and a variable number of beta distributions to account for the clonal and subclonal peaks.

We ran MOBSTER using the default parameters, except employing a minimum 5% VAF threshold and lowering the minimum number of mutations to compose a cluster to 5 in WES samples due to the low number of mutations. We then manually QC’d all 377 WES samples and 10 WGS, tuning the fitting parameters to better represent the data (for instance, when the clonal peak had been called at a low frequency despite the median tumour purity being 95%).

### Phylogenetic inference of longitudinal methylation data

A novel Bayesian phylogenetic method was used to reconstruct the evolutionary relationships and the time to most recent common ancestor (tMRCA) of longitudinal samples from the same patients. This was carried out in the BEAST v.1.8.4 framework (Drummond and Rambaut 2007; Drummond et al. 2012) using custom models implemented in PISCA v1.1 (available from https://github.com/adamallo/PISCA) (Martinez et al. 2018).

EVOFLUx provided an estimate of the age of the patient when the MRCA of each bulk sample emerged. To estimate the methylation status of each fCpG at the sample’s MRCA in each of our longitudinal samples, we discretised the fCpGs as described above (RNAseq Methods).

We implemented a 4-parameter biallelic binary substitution model analogous to the pre- growth EVOFLUx model in PISCA. This plugin contains all the required statistical machinery to use this model for somatic phylogenetic estimation. The biallelic binary substitution model has three relative rate parameters 1) heterozygous methylation 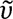, 2) homozygous demethylation 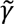, and 3) heterozygous demethylation 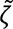, where homozygous methylation 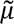 was normalised to 1. For all relative transition rate parameters, a lognormal prior with mean 1 and standard deviation of 0.6 was used, with a half normal prior with mean 0 and standard deviation 0.13 for the molecular clock rate, using a strict clock model for the rate of evolution across the tree. Two demographic tree models, constant population size (Kingman 1982) and exponential growth (Griffiths and Tavaré 1994), were compared by marginal likelihood estimation using path-sampling (Baele et al. 2012) and a constant population model was deemed more appropriate.

MCMC chains were run for 100 million generations sampled every 100,000 generations and convergence was assessed using Tracer v.1.7 (Rambaut et al. 2018), ensuring effective sample sizes greater than 500 for all parameters. Maximum clade credibility trees were then made using 10% burn-in and medium node heights. The resulting trees were plotted using ggtree (Yu et al. 2017).

### Phylogenetic inference of SNVs from WGS data

Each bulk sample is represented by a set of clonal mutations found during the deconvolution of WGS data (see above). Where a mutation was deemed absent in the clonal peak, the reference nucleotide was used. Mutational signature assignment (Díaz-Gay et al. 2023) was used to select mutations in the clock-like SBS1 channel (Tate et al. 2019). BEAST v.1.10 (Suchard et al. 2018) was then used with the simple binary substitution model (since SBS1 effectively represents just C to T substitutions), a strict clock model, a constant population size prior (Kingman 1982), and a flat prior on the age of most recent common ancestor (from zero to earliest patient sample), with ancestral state estimation at the root. Chains were run and ESS values assessed as described above. The distance between the ancestral state of the root at each MCMC state, and the clock rate were used to calculate the expected evolution distance between the root and the known germline. This was used to inform the length of the branch between germline (at birth) and the MRCA of the samples.

### Survival Analysis

Clinical analyses were performed in CLL for time to first treatment (TTFT) and overall survival (OS) from the time of sampling. Tumour growth rate (*θ*), effective population size (*N*_e_) and epigenetic switching rates were analysed as continuous variables in univariate Cox regression models for both TTFT and OS. The effect size of hazard ratios (HRs) for each evolutionary variable were analysed considering different scaling factors. In particular, the growth rate was analysed in exponential growth per year (i.e., for *θ = 1*, the population per year is e=2.71 times bigger) ; the *N*_e_ was considered per million cells; the cancer age or time from the MRCA was analysed for each 10 years; the homozygous to heterozygous (de)methylation rates (γ and *µ*, respectively) and heterozygous to homozygous methylation rate (*v*) were analysed multiplied by a factor of 100 (per allele per year); and heterozygous to homozygous demethylation rate (ζ) was multiplied by a factor of 1,000. In addition, growth rate and effective population were analysed as continuous variables in multivariate Cox regression models together with *TP53* aberrations (considering mutations and deletions together), IGHV gene mutational status and the age of patients at sampling. Kaplan-Meier (KM) curves were generated for low and high growth rate and effective population size within IGHV subtypes using maximally selected log-rank statistic using maxstats package (version 0.7-25). *P* values from KM curves were derived using the Log-rank statistic. Survival (version 3.5-7), survminer (version 0.4.9) and ggsurvfit (version 0.3.1) packages were used under R version 4.3.1.

## Data availability

No new data was generated in the course of this study. The harmonised and filtered methylation matrix is available (preferred browser Firefox) at: http://inbcg.bsc.es/hcli_priv/publications_data/fluctuating_cpgs_gabbutt_duran_ferrer_2023/QC_2204_ssNob.tsv.7z - the password for this file is available upon request to the authors.

Previously published DNA methylation data re-analysed in this study can be found under accession codes: B cells, EGAS00001001196; ALL, GSE56602, GSE49032, GSE76585, GSE69229; MCL, EGAS00001001637, EGAS00001004165; CLL, EGAD00010000871, EGAD00010000948, EGAD00010001975; MM, EGAS00001000841; DLBCL, EGAD00010001974.

CLL gene expression data is available EGAS00001000374 and EGAS00001001306. ChIP-seq datasets are available from Blueprint https://www.blueprint-epigenome.eu/ under the accession EGAS00001000326.

Matched WES and WGS are available under accessions EGAS00000000092 and EGAD00001008954 respectively.

## Code Availability

The EVOFLUx code employed to infer the evolutionary history of cancer samples from methylation array data is attached as a bundled Zip file and will be made publicly available via Github prior to publication at https://github.com/CalumGabbutt. Further code will also be made available at https://github.com/Duran-FerrerM and https://github.com/adamallo/PISCA.

## Ethics statement

The study was approved by the ethics committees of the institutions that generated the data used in the present study.

