## Supplementary Information for "Evolutionary dynamics of 1,976 lymphoid malignancies predict clinical outcome"

### Supplementary Figures

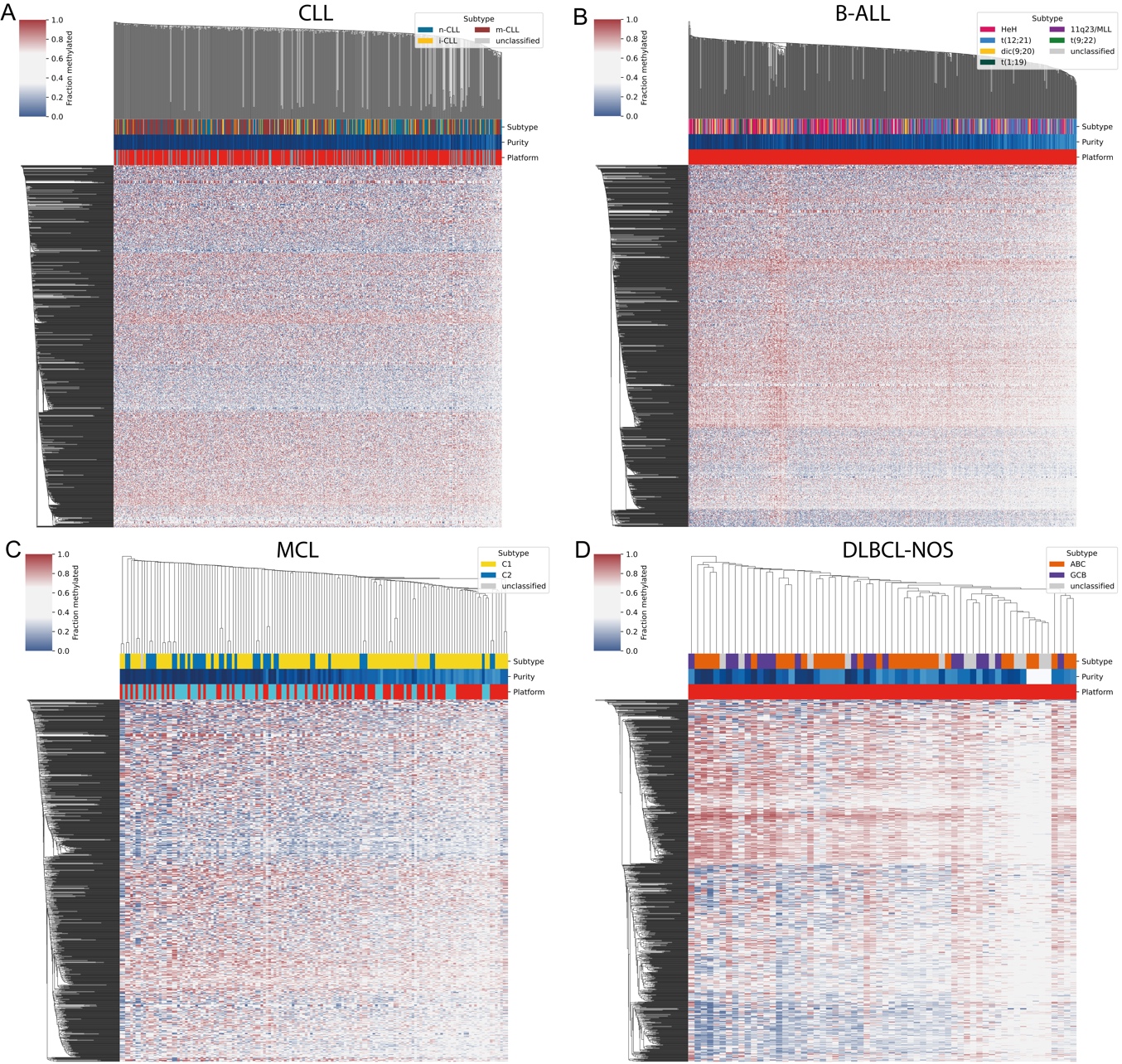

**Supplementary Figure 1 – Heatmaps of putative fCpG loci by disease**

Clustered heatmaps (hierarchical with average linkage and a Euclidean metric) of our set of 978 pan-lymphoid fCpGs with disease-specific subtypes annotated in: **A** chronic lymphocytic leukaemia (CLL), **B** B-cell acute lymphoblastic leukaemia (B-ALL), **C** mantle cell lymphoma (MCL), and **D** diffuse large B-cell lymphoma - not otherwise specified (DLBCL-NOS).

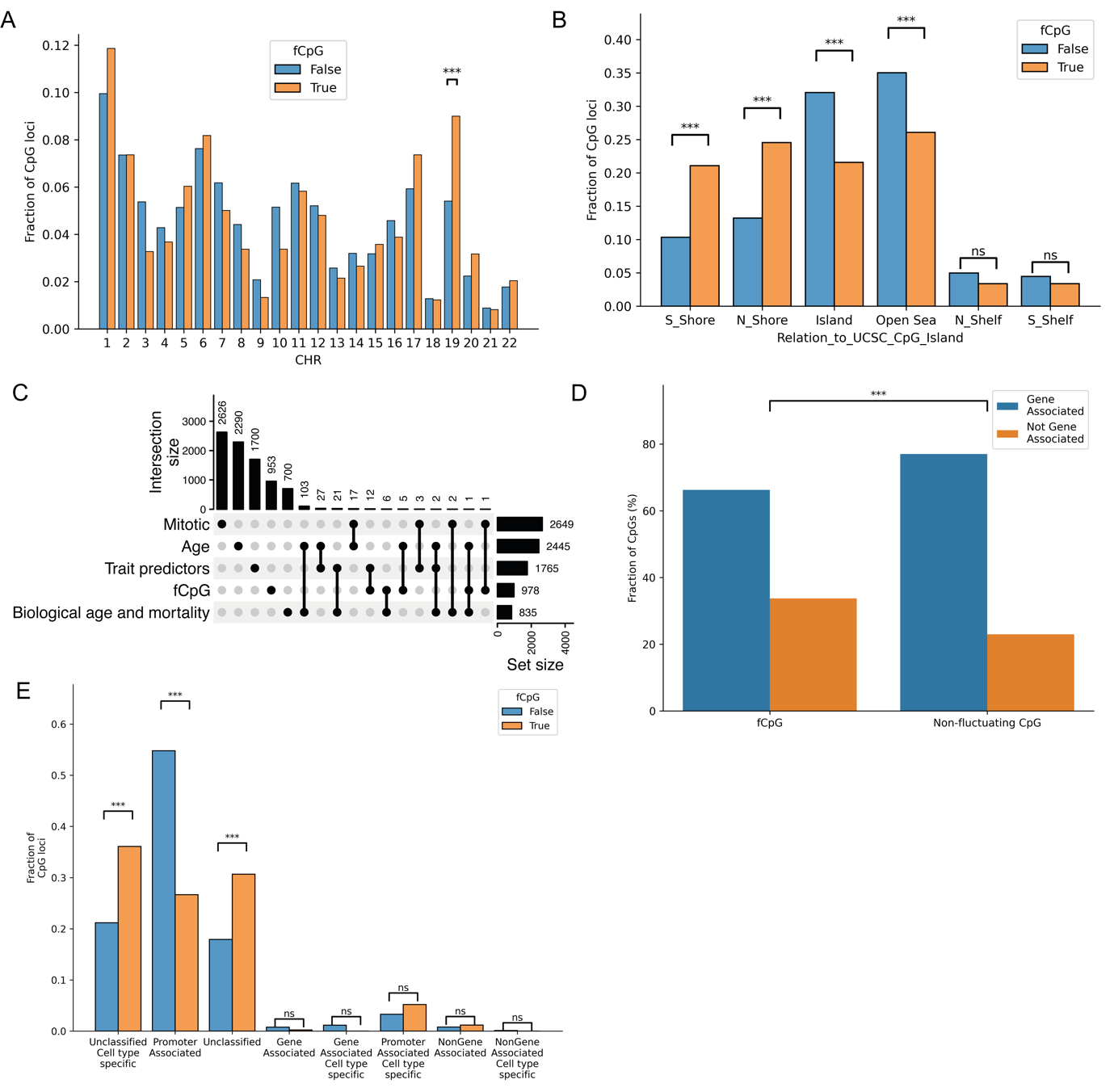

**Supplementary Figure 2 – Genomic properties of fCpG loci**

**A:** fCpG loci are similarly distributed across the genome when compared to non-fCpG loci also on the 450K Illumina array, except a significant enrichment on chromosome 19 (p=2.2e-5, pairwise $\chi^{2}$ tests, holm-sidak (hs) correction). **B:** Paired bar charts demonstrating that fCpG loci are enriched on the shores of CpG Islands but depleted on both the islands themselves and in the Open Sea. **C:** A comparison of the mutual overlap between different epigenetic clocks: mitotic (Duran-Ferrer et al. 2020; Yang et al. 2016; Teschendorff 2020; Youn and Wang 2018; Zhou et al. 2018), chronological age (Bocklandt et al. 2011; Garagnani et al. 2012; Hannum et al. 2013; Horvath 2013; Lin et al. 2016; Vidal-Bralo, Lopez-Golan, and Gonzalez 2016; Weidner et al. 2014; Zhang et al. 2019; Horvath et al. 2018; Shireby et al. 2020), gestational age (Bohlin et al. 2016; Knight et al. 2016; Lee et al. 2019; Mayne et al. 2017; McEwen et al. 2020), biological age and mortality (Belsky et al. 2022; Levine et al. 2018; Lu et al. 2019), and trait predictors (McCartney et al. 2018; Liang et al. 2020) and fCpGs. **D:** fCpG loci are significantly less likely to be associated with a gene (annotations provided by UCSC, p<0.001, $\chi^{2}$ test). **E:** fCpG loci are significantly less likely to be associated with a promotor, and much more likely to be either unclassified or unclassified but associated with a specific cell type (annotations provided by ENCODE (Dunham et al. 2012)) (all significant p values < 10^-10^, pairwise $\chi^{2}$ tests, hs correction).

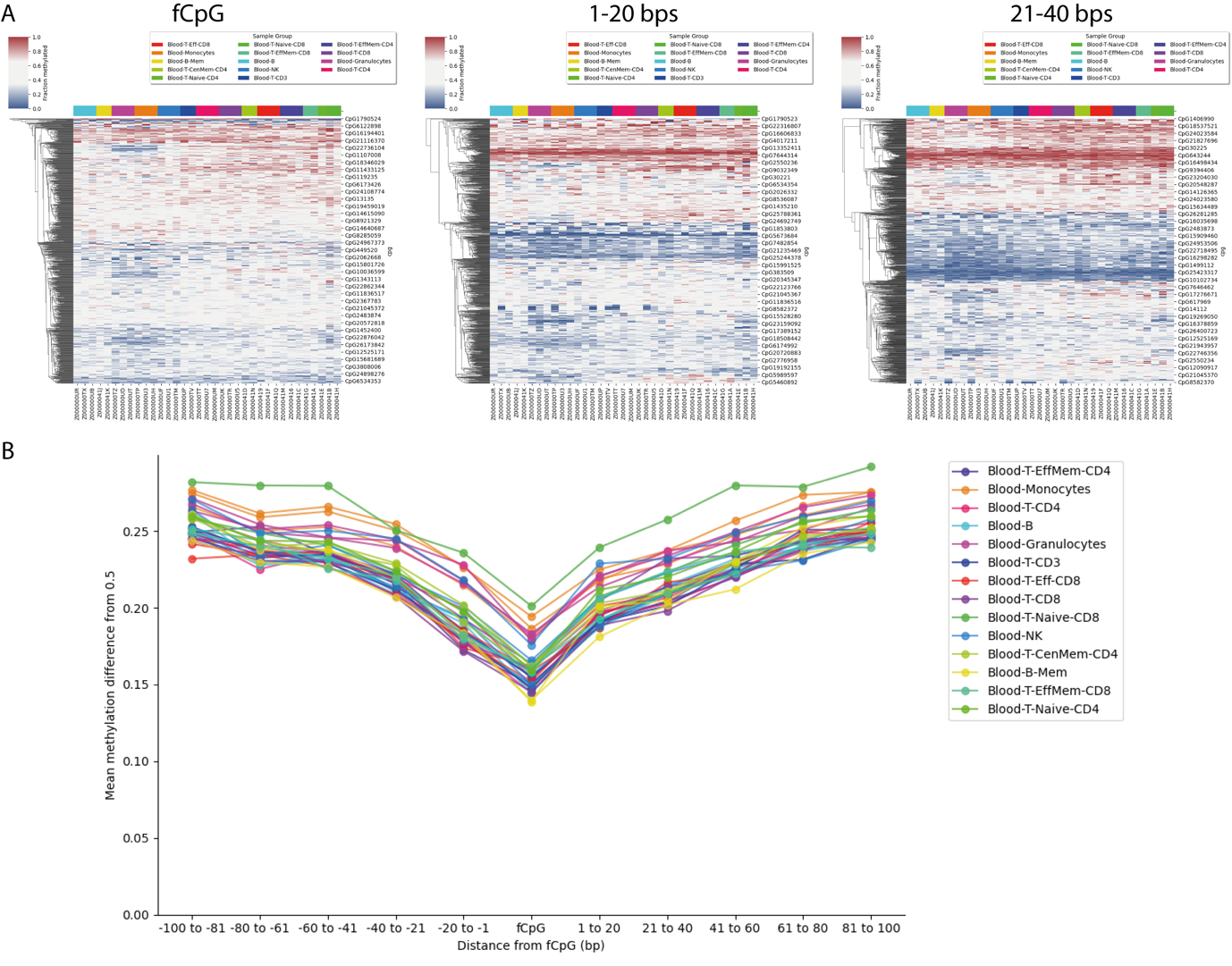

**Supplementary Figure 3 – Local genomic neighbourhood surrounding fCpG loci**

**A:** Heatmaps of the methylation values of sorted healthy B- and T-cells (Loyfer et al. 2023) of fCpG loci, CpG loci within 1-20 bps and 21-40 bps. In polyclonal populations, measurements of bulk sorted population are expected to have intermediate methylations, whereas CpG loci under strict regulation are expected to be hypo- or hypermethylated. **B:** The average absolute difference from 0.5 for methylation values from fCpG loci and CpG loci in the local neighbourhood, in different B- and T- cell populations. The fraction of hypo- and hyper- methylated CpG loci increases as a function of distance from the reference fCpG locus.

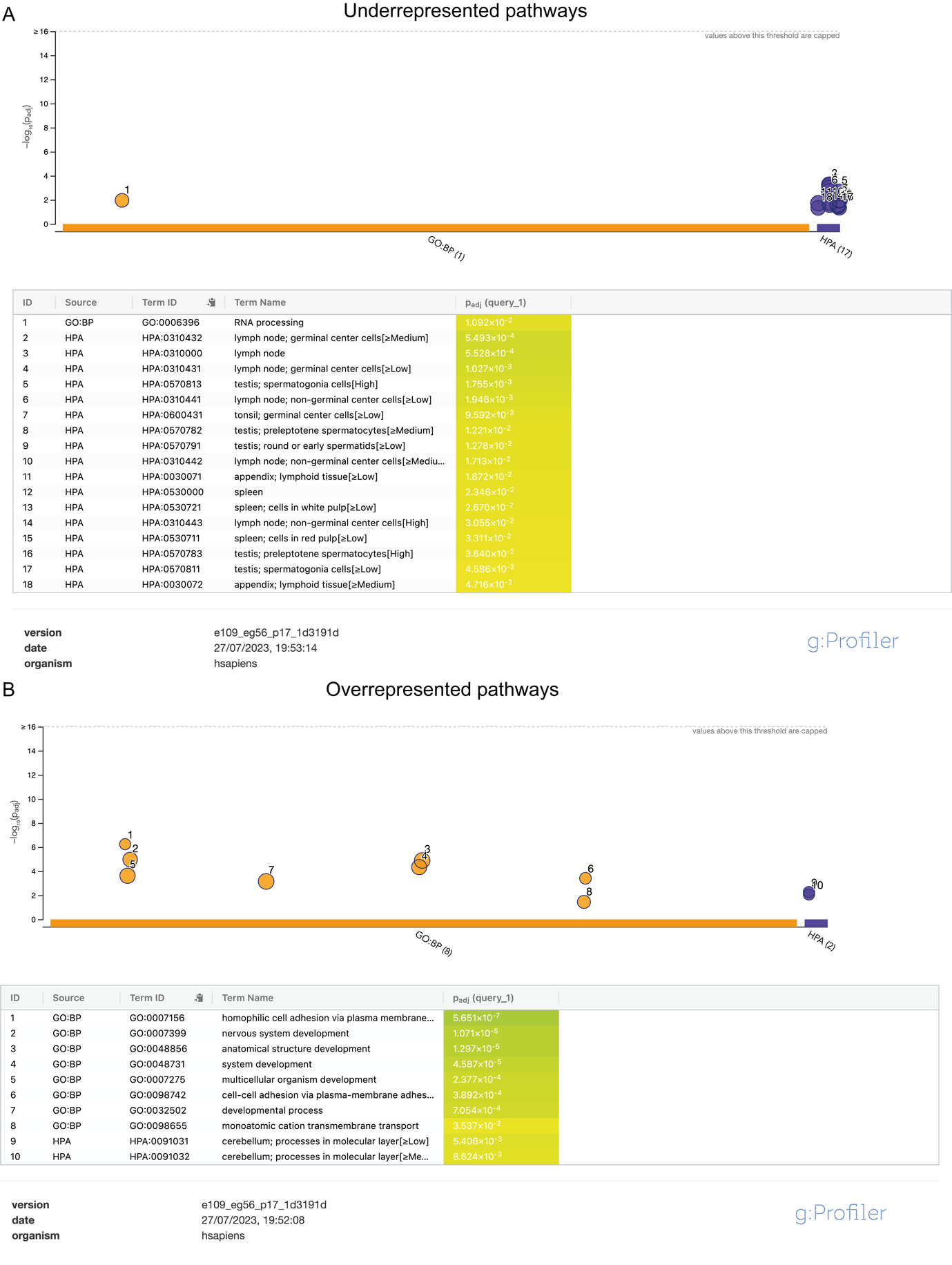

**Supplementary Figure 4 – Gene-set enrichment analysis of fCpG-associated genes**

Gene-set depletion (**A**) and enrichment (**B**) analysis of genes associated with fCpG loci according to UCSC [ref] using gProfiler, with the target gene-space corresponding to all genes with an associated CpG locus present on the 450K Illumina methylation bead array. The Gene Ontology biological processes (Ashburner et al. 2000) and the Human Protein Atlas (Uhlén et al. 2015) databases were employed. fCpG associated genes were significantly enriched in developmental pathways but depleted in lymph, testis, and tonsil tissues.

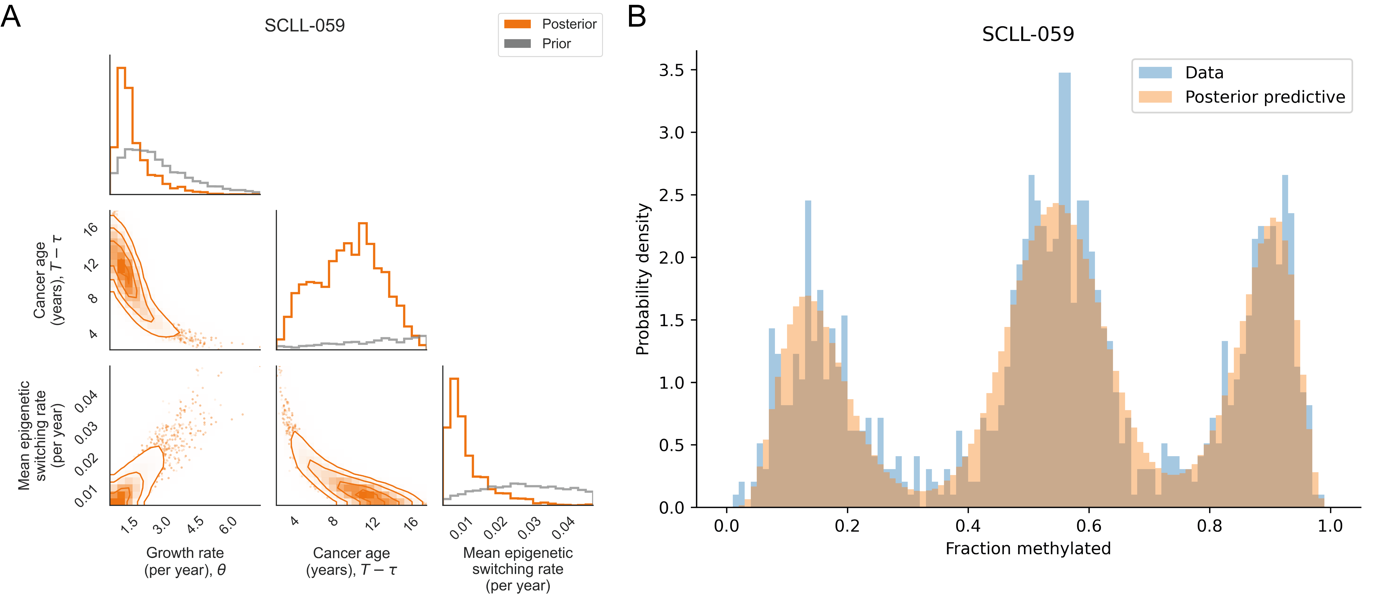

**Supplementary Figure 5 – Example EVOFLUx posterior and fit**

**A:** An example of the posterior resulting from running EVOFLUx on a CLL sample – a pairs plot showing the marginal (diagonal) and pairwise (off-diagonal) inferred posterior (orange) distributions, with the prior distributions overlaid (gray). The posteriors show marked tightening compared to the priors, demonstrating the parameters are well informed by the data. **B:** A histogram of the fCpG methylation distribution (blue) with the posterior predictive of the model fit overlaid (orange).

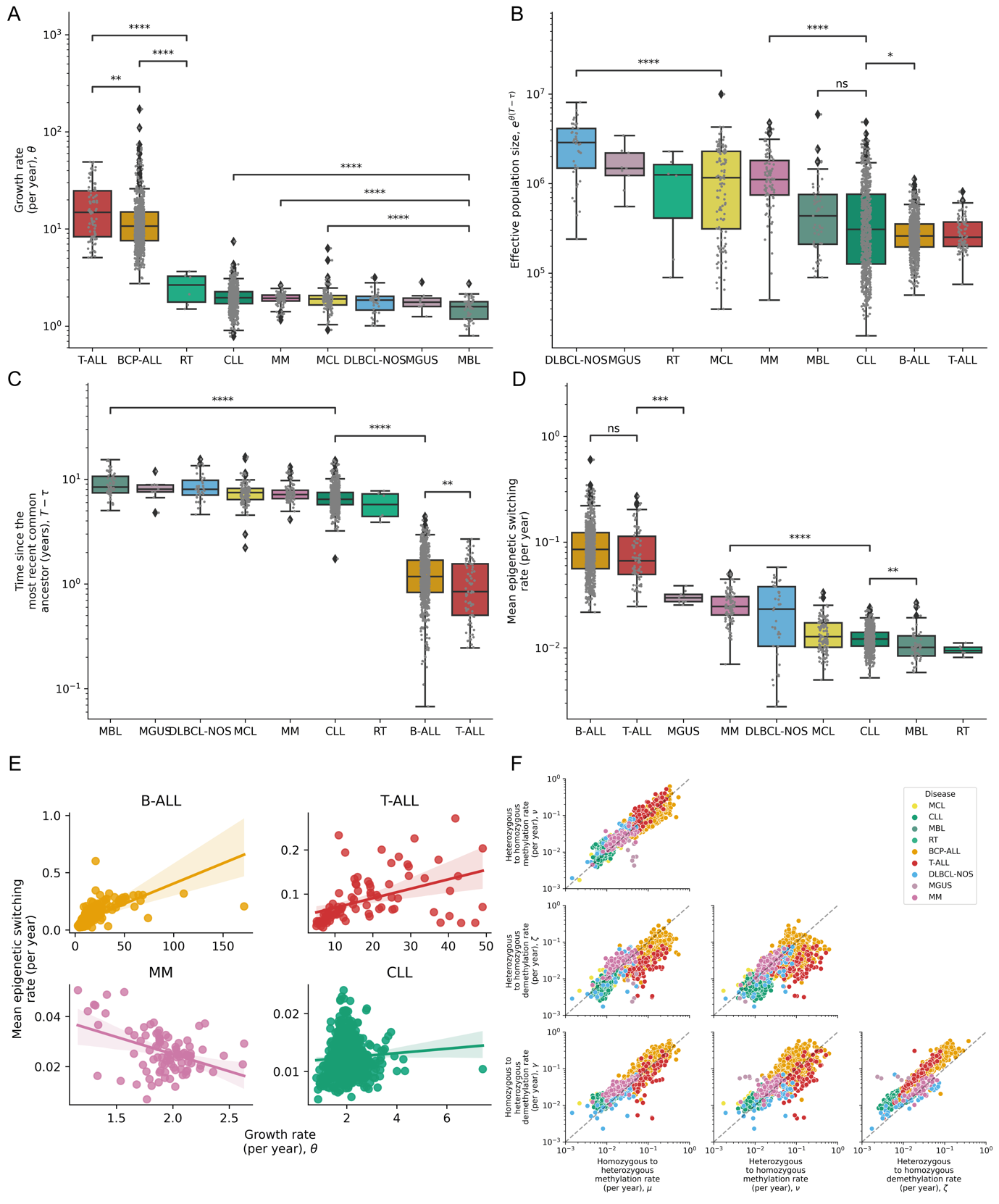

**Supplementary Figure 6 – EVOFLUx captures lymphoid cancers’ evolutionary histories and methylation epimutation dynamics**

**A-D:** Boxplots (whiskers extending to ±1.5×IQR) showing the distribution of inferred growth rate (**A**), effective population size (**B**), time since the most recent common ancestor (**C**), and mean epigenetic switching rate (**D**) by disease. For interpretability, only a subset of the pairwise p values are annotated (Mann-Whitney U tests, hs correction). **E:** Linear regression between the growth and epigenetic switching rates separated by cancer types. There is a positive association in B-ALL (p= 2.4e-98, R^2^=0.44) and T-ALL (p=5.9e-06, R^2^=0.22), a weak negative association in MM (p=1.6e-05, R^2^=0.18) and no association in CLL (p=0.060, R^2^=0.005) or the other entities. **F:** A pairs plot comparing the four epigenetic switching rates (μ, υ, γ and ζ) in each of the cancers, coloured by disease.

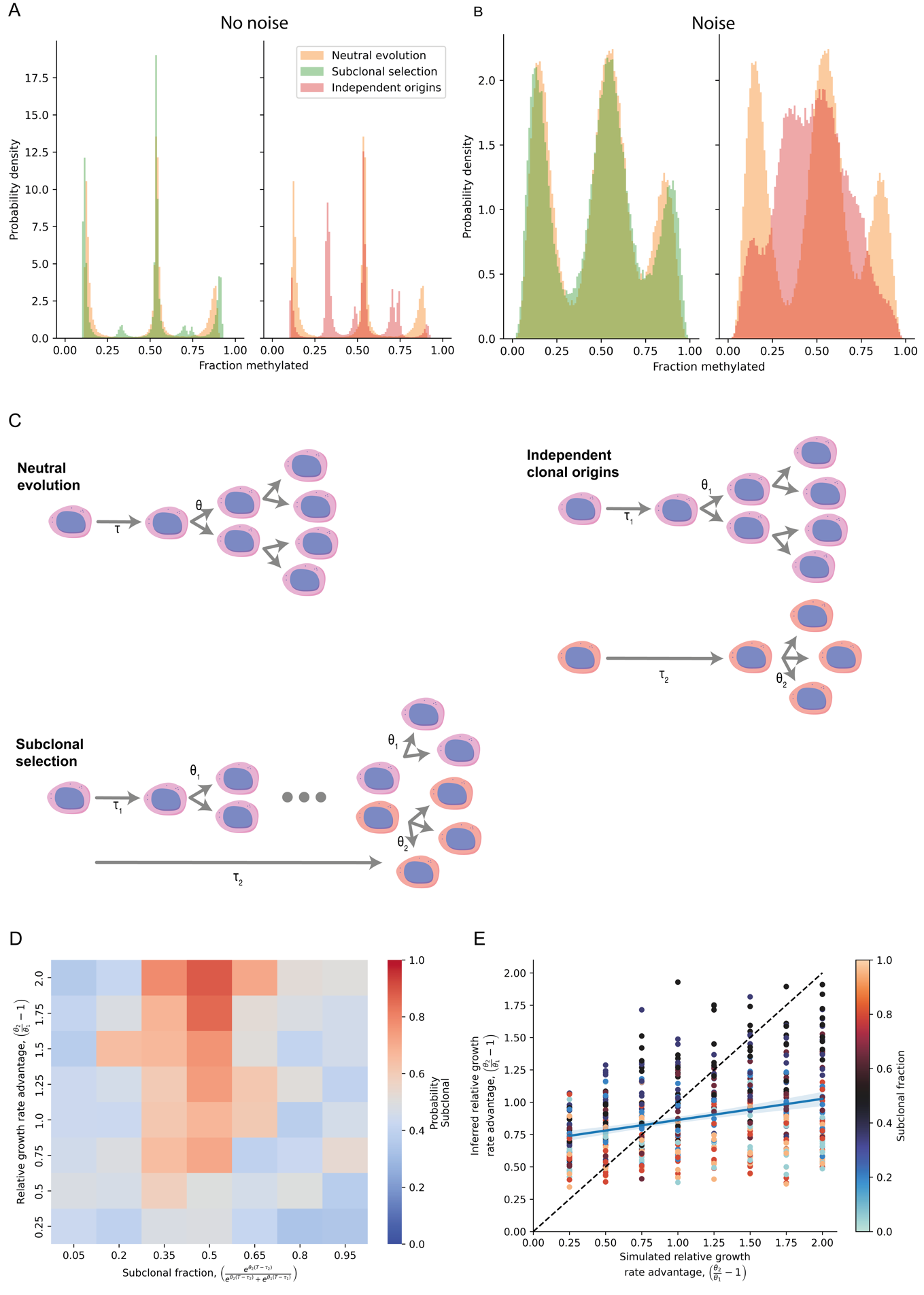

**Supplementary Figure 7 – Simulations of non-neutral evolutionary dynamics**

**A-B:** Simulations comparing the fCpG methylation distribution in the neutral (orange) case vs subclonal selection (left, green) and independent clonal origins (right, red) in the case with no noise (**A**) and with noise added (**B**). **C:** Illustrations of three alternative evolutionary models: neutral evolution, in which the cancer population with a MRCA emerging at time $\tau$ grows exponentially at rate $\theta$; subclonal selection, in which an initial population emerging at time $\tau_{1}$ growing at rate $\theta_{1}$ is outcompeted by a fitter subclonal emerging at rate $\tau_{2}$ with growth rate $\theta_{2}$; and independent clonal origins, in which the fitter clone emerging at time $\tau_{2}$ with growth rate $\theta_{2}$bears no clonal relationship to the initial clone. **D:** A heatmap of showing how the subclonal model weight of a simulation varies with the fraction of the population made up of the fitter subclone and the growth advantage of the fitter subclone. **E:** A comparison of the inferred relative growth advantage of a simulation vs the actual ground truth value, where the colour of the point reflects the fraction of the subclonal population.

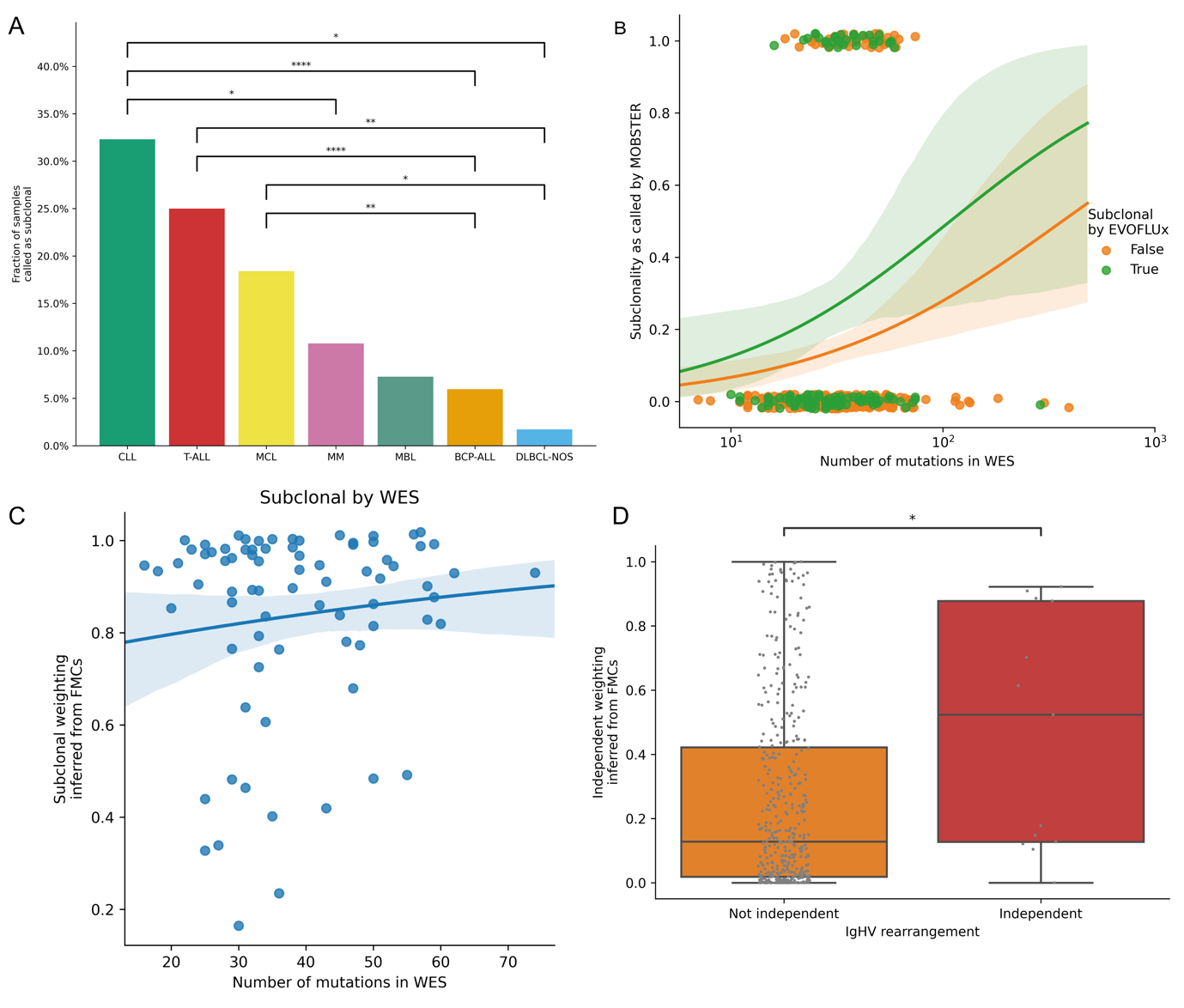

**Supplementary Figure 8 – Inference and validation of subclonal selection using fCpG loci**

**A:** Bar chart comparing the fraction of cancer samples identified as subclonal by EVOFLUx across disease (pairwise $\chi^{2}$ tests, holm-sidak (hs) correction). **B:** Logistic regression between the probability of a cancer being identified as subclonal via running our subclonal deconvolution tool, MOBSTER, on WES data and the number of mutations detected in the sample. Regression run separately for samples also identified as subclonal/neutral via EVOFLUx. **C:** Logistic regression between the inferred subclonal weighting by EVOFLUx and the number of mutations detected in WES finds no association between the two (p>0.05, logistic regression). **D:** Boxplots comparing the distribution of independent model weightings inferred by EVOFLUx in samples containing multiple IgHV rearrangements (likely independent cancers) vs those with only a single IgHV rearrangement (likely a single clonal origin).

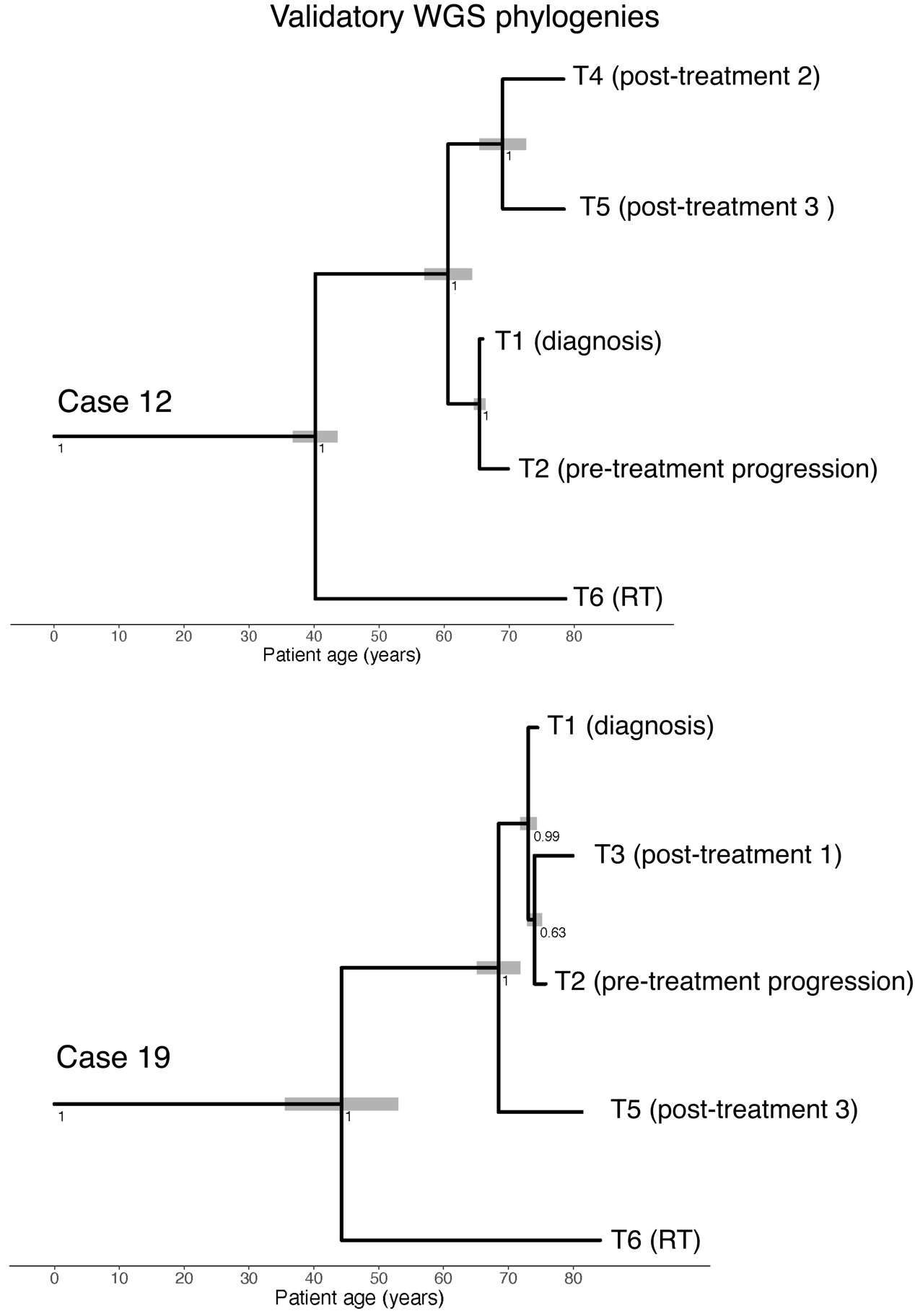

**Supplementary Figure 9 – WGS SNV phylogenies validate fCpG phylogenies in CLL**

Phylogenies reconstructed on matched WGS SNV data have a similar topology and absolute timing of branch points as to those reconstructed using fCpGs (fig 4).

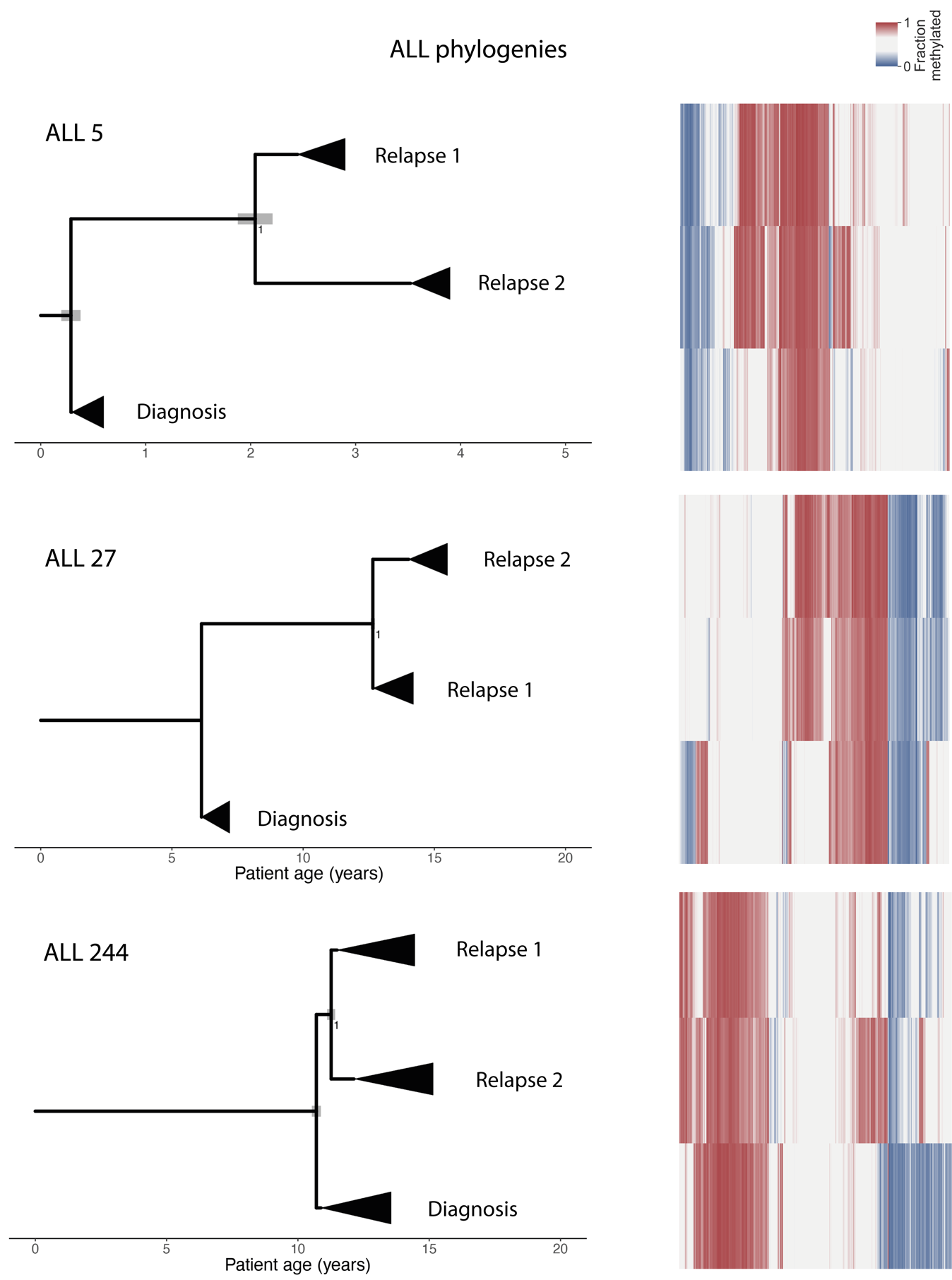

**Supplementary Figure 10 – fCpG phylogenies in ALL**

(left) The reconstructed phylogenies of the relationship between samples collected longitudinally in individual ALL patients, annotated with the clinical classification of each sample. The black triangles represent the time that occurred since the most recent common ancestor, taken as the posterior median of $T-\tau$ from the single-sample EVOFLUx inferences. (right) A heatmap representing the 978 fCpG loci, with the colour a representing the fraction methylated (0% blue, 100% red).

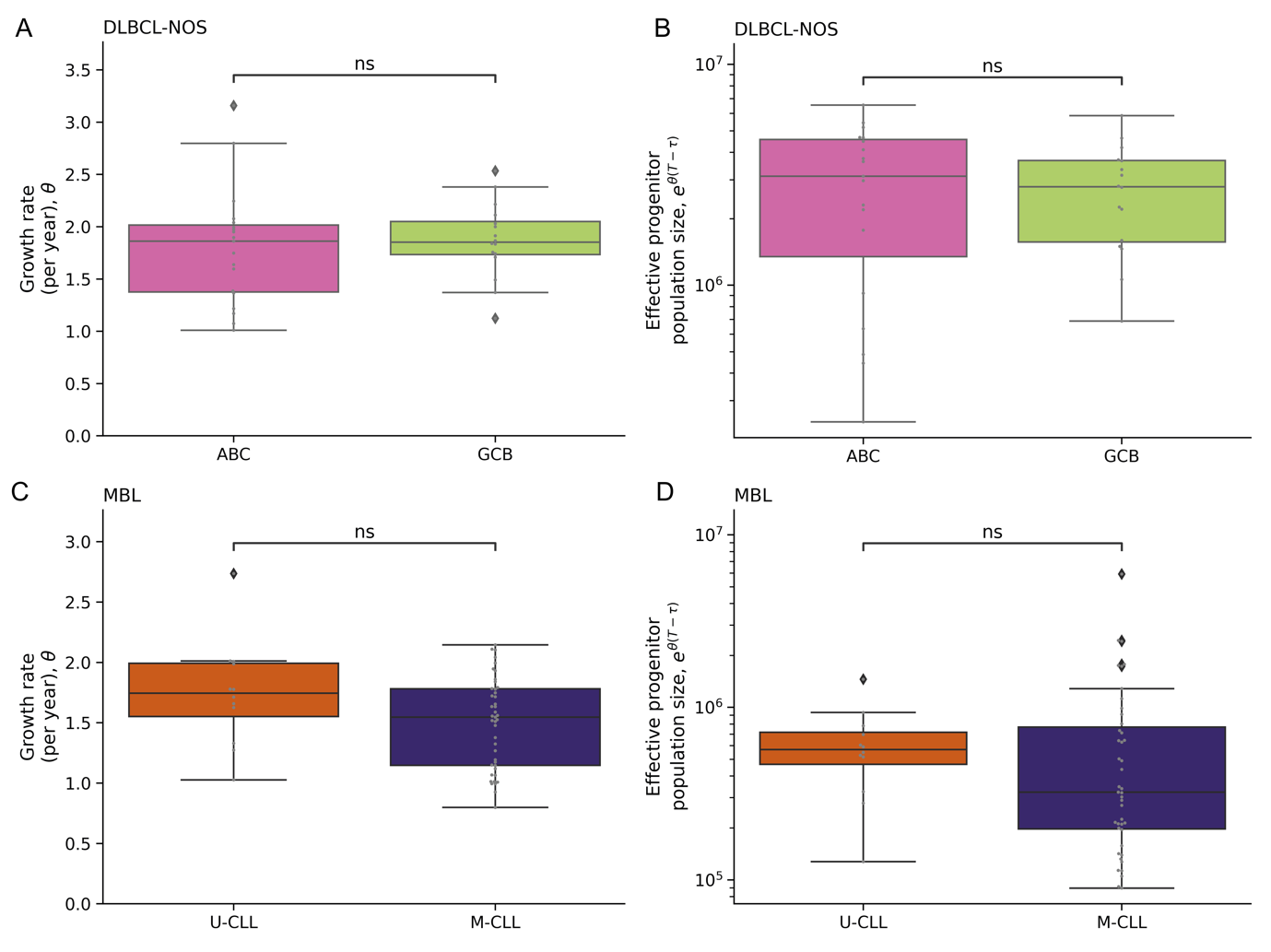

**Supplementary Figure 11 – Additional subtype comparisons of the inferred evolutionary parameters**

The inferred growth rate (**A&C**) and effective population size (**B&D**) of individual cancer samples separated by molecular subtype in diffuse large B-cell lymphoma (**A&B**, DLBCL-NOS) and monoclonal B-cell lymphocytosis (**C&D**, MBL). Differences between subtypes were tested using Mann-Whitney U tests.

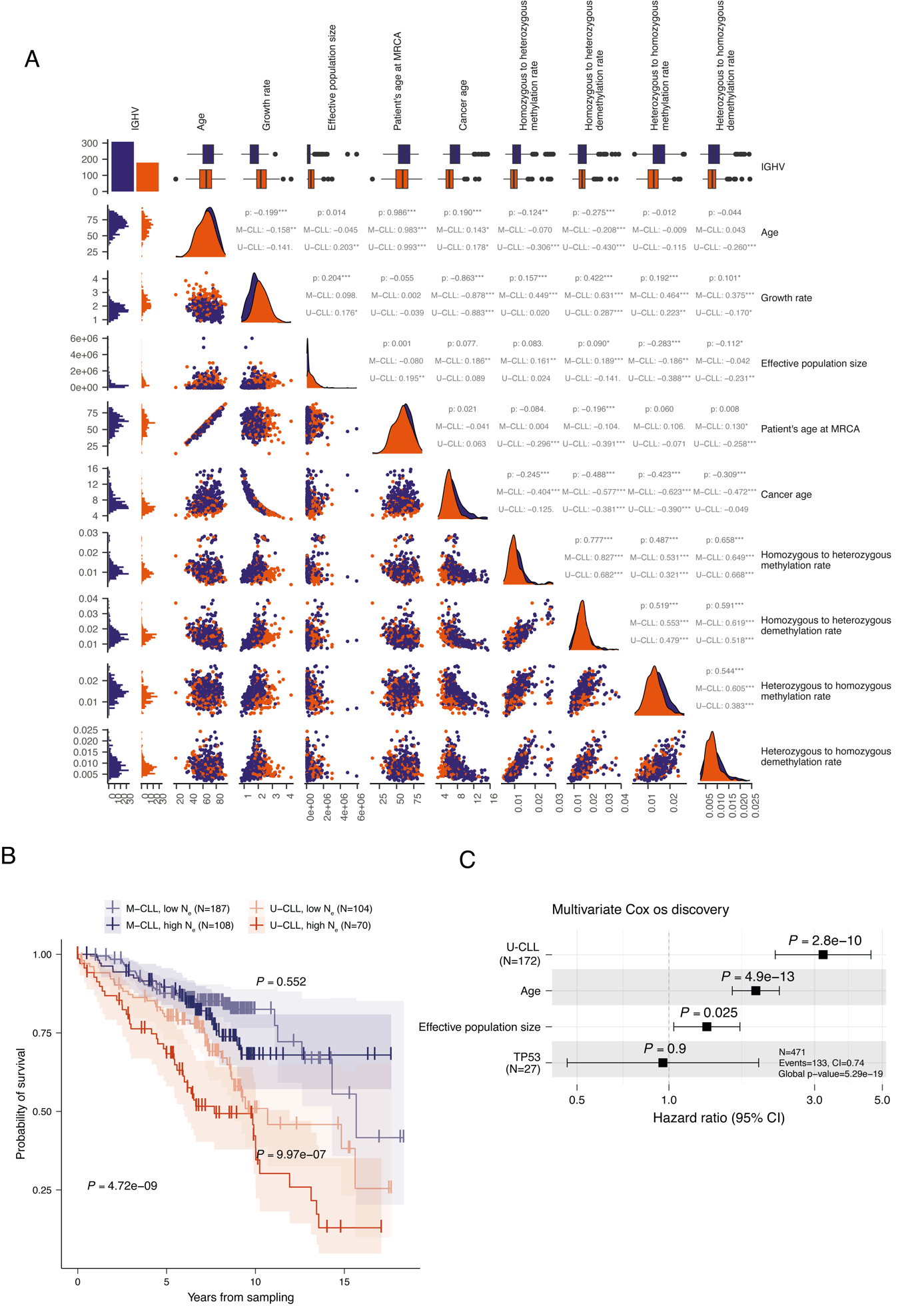

**Supplementary Figure 12 – Correlations in CLL evolutionary parameters and additional survival analysis**

**A:** A pairs plot showing the marginal relationships between the observed evolutionary parameters in the discovery cohort as scatter plots (lower left triangle), histograms (leading diagonal) and summary correlation coefficients (stars indicate significance). Patients separated by IGHV mutational status (unmutated orange, mutated purple). **B:** Kaplan-Meier curves comparing the OS between patients with high vs low (cut-off values identified using maxstat statistics) inferred effective population sizes (N_e_), separated by IGHV mutational status. **C:** Multivariate Cox regression of the OS shows the N_e_ is significant when controlling for IGHV status, TP53 status and age in the discovery cohort.

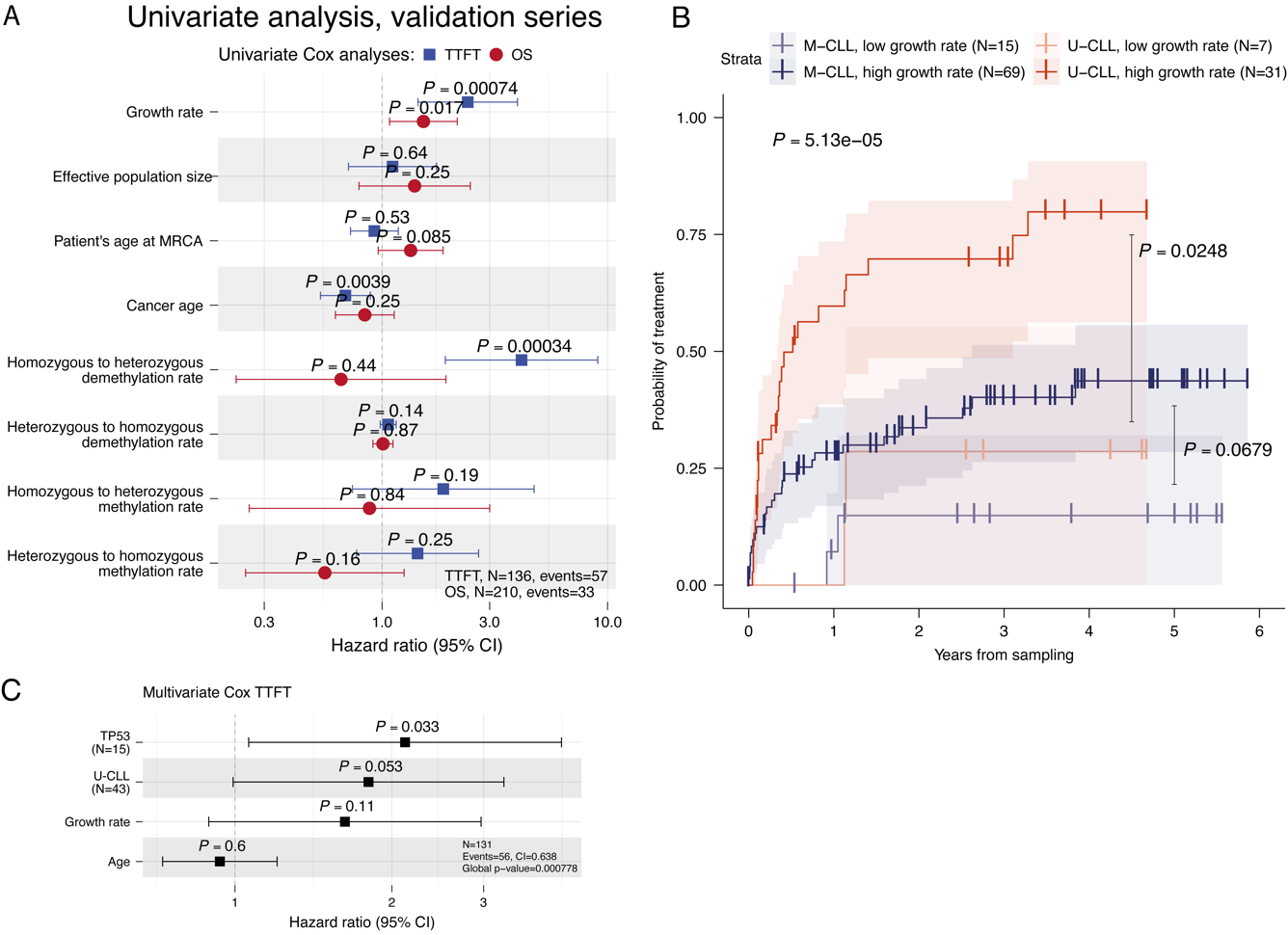

**Supplementary Figure 13 – Survival analysis in validation cohort**

**A:** Univariate survival analysis of the time to first treatment (TTFT, blue) and overall survival (OS, red) in the validation CLL cohort for evolutionary variables inferred via EVOFLUx. Note this cohort contains a mixture of treated and untreated samples, of which only the untreated samples were included in the TTFT analysis. **B:** Kaplan-Meier curves comparing the TTFT between patients with high vs low (cut-off values identified using maxstat statistics) inferred cancer growth rates in the validation cohort, separated by IGHV mutational status. **C:** Multivariate Cox regression of the effect of the cancer growth rate on the in the validation cohort, controlling for IGHV status, TP53 status and age.

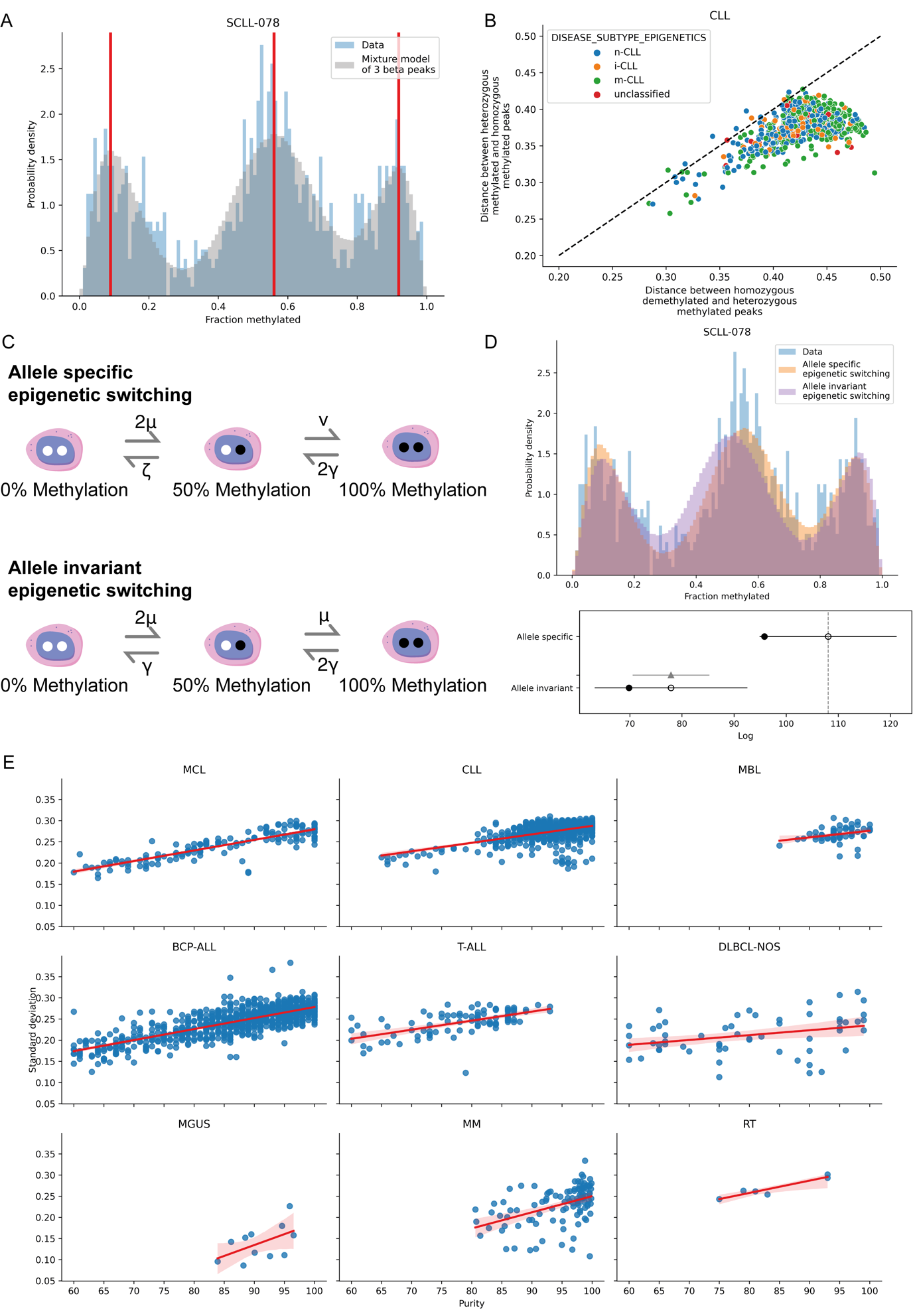

**Supplementary Figure 14 – Allele-specific epigenetic switching**

**A:** A histogram displaying the fCpG methylation value distribution of a CLL sample (blue) with a posterior predictive of a beta mixture model (with 3 components) overlaid (gray). The vertical red lines indicate the mode of the 3 inferred beta mixture components. **B:** A scatterplot comparing the distance between the inferred modes of the two homozygous peaks and the heterozygous peak in the CLL cohort. The distance between the homozygous unmethylated and the heterozygous peak is larger in the vast majority of samples. **C:** A schematic illustrating two competing models, one in which the methylation of a given allele depends on the methylation status of the other (top, 4 parameters) and a model in which the two alleles are independent (bottom, 2 parameters). **D:** (top) A histogram displaying the fCpG methylation value distribution of a CLL sample (blue) with the model fits of the allele specific (orange) and allele invariant epigenetic (lilac) switching models overlaid. (bottom) A leave one out cross validation model comparison, showing the allele specific model is strongly preferred. **E:** Scatterplots showing the association between tumour purity and the standard deviation of fCpG methylation in lymphoid cancers.

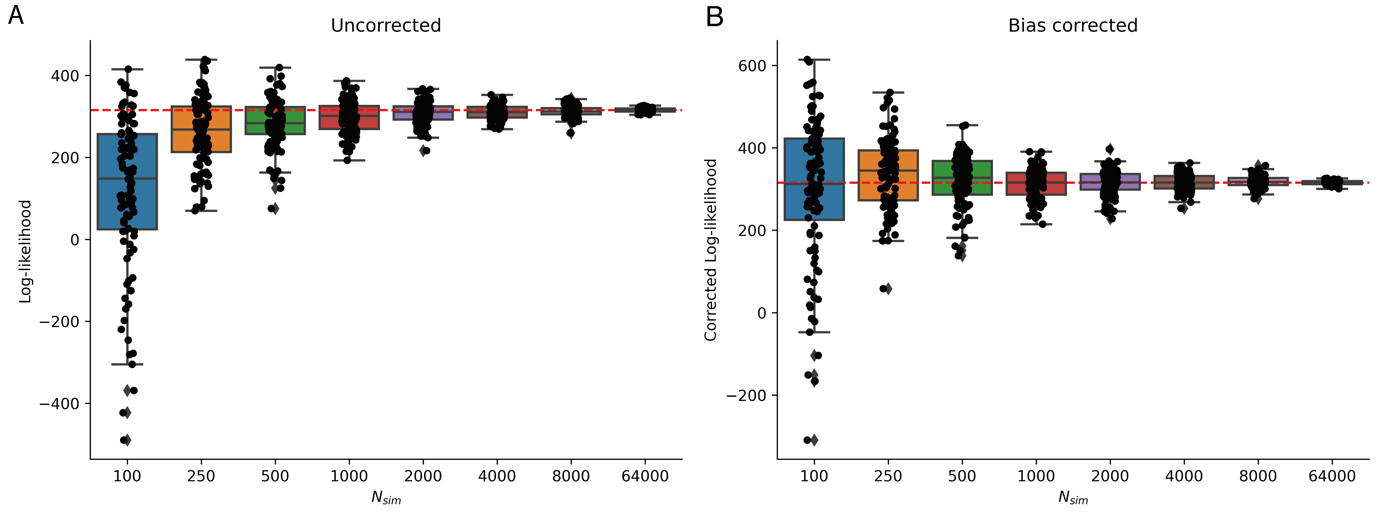

**Supplementary Figure 15 – Loglikelihood correction**

**A:** Repeated calculations (n=100) of the uncorrected synthetic loglikelihood (LL) value of simulated data calculated at the parameter values used to simulate the data, as a function of the number of simulations (n_sim_) used in the LL calculation. Using a finite n_sim_ systematically underestimates the true LL. **B:** The bias corrected synthetic LL, the mean of which does not vary as a function of n_sim_.

### EVOFLUx – a novel method to simulate and infer the evolutionary history of well-mixed cancers from methylation-based lineage markers

In our previous work (Gabbutt et al. 2022), an analytic hidden Markov model was derived to describe the distribution of fluctuating methylation clocks in a fixed effective population of $N_{e}$ cells, replacing each other at a rate $\lambda$ per stem cell and with each fCpG loci allowed to alter its methylation state at a methylation/demethylation rate $\mu$/$\gamma$ respectively. In principle, the mathematics developed there applies for all values possible cell numbers; however, the number of possible states (i.e., the different possible combinations of heterozygous and homozygous (de)methylated cells at a specific CpG locus) grows as $\sim{N_{e}}^{2}$, hence this approach is inappropriate for large populations of cells. Furthermore, the previous approach assumed a fixed number of cells, rather than a growing population.

Hence, to extend our analysis to large, growing cell populations, such as those found within a blood cancer, an alternative approach to modelling the probability distribution that does not scale with the number of stem cells must be taken. Instead, we took a stochastic approach to approximate the fCpG methylation distribution in an arbitrarily large, exponentially growing population. Rather than considering the individual states and the possible flows between them, we constructed equivalent stochastic jump differential equations. Finally, whereas previously it was the stochastic replacement of one stem cell by another that counteracted the ongoing (de)methylation, effectively resynchronising the methylation clocks upon a random clonal expansion; in the case of a growing cancer this neutral drift is negligible when compared to the exponential growth of each lineage within the tumour.

In summary, then, our model consists of a single cell that undergoes random (de)methylation until time $\tau$, at which point it begins exponentially growing at rate $\theta$ per year until the patient’s age at the time of sampling, $T$, is reached. We allow for the possibility of allele specific methylation, and therefore require four epigenetic switching parameters to describe the system (fig 2A):

- $\mu$ – Homozygous to heterozygous methylation rate (per year)
- $\upsilon$ – Heterozygous to homozygous methylation rate (per year)
- $\gamma$ - Homozygous to heterozygous demethylation rate (per year)
- $\zeta$ – Heterozygous to homozygous demethylation rate (per year)

We employed four separate methylation switching parameters as these were necessary to reproduce the observed data. In CLL patient data, the distance between the homozygous unmethylated peak and the heterozygous methylated peak was almost always lower than the distance between the homozygous methylated peak and the heterozygous methylated peak (fig. S14A&B). A model where the homozygous to heterozygous rate is the same as the heterozygous to homozygous rate (i.e. $\upsilon=\mu$ and $\zeta=\gamma$, fig. S14A) provided a significantly worse fit to the data (accounting for the higher degrees of freedom in the 4-parameter model) than one in which the parameters are free to vary (fig. S14D).

We assume that each methylation site is equally likely to be homozygous methylated or unmethylated at $t=0$, as in our previous work. We shall use the notation that there are $m$ cells with both alleles methylated, $k$ cells with one allele methylated and $w$ cells with neither allele methylated. To calculate the methylation probability distribution of the single transformed cell at $t=\tau$, the system is evolved according to the following set of differential equations:

$$\frac{d}{dt}\left( \begin{matrix} p\left( m;t \right) \\ p\left( k;t \right) \\ p\left( w;t \right) \end{matrix} \right)=\left( \begin{matrix} -2\gamma& \upsilon& 0 \\ 2\gamma& -\left( \zeta+\upsilon\right) & 2\mu\\ 0 & \zeta& -2\mu\end{matrix} \right)\left( \begin{matrix} p\left( m;t \right) \\ p\left( k;t \right) \\ p\left( w;t \right) \end{matrix} \right)$$

To calculate the initial state of the transformed cell at a particular locus at $t=\tau$, the state $\left( m, k, w \right)$ is drawn from a categorical distribution with probabilities $\left( p\left( m;t=\tau\right), p\left( k;t=\tau\right), p\left( w;t=\tau\right) \right)$, which may be calculated using matrix exponentiation. The subsequent evolution of the system for $t>\tau$ is then modelled using discrete time stochastic jump equations with a time step, $\delta t$, that is sufficiently small that the probability of double jumps is negligible:

$$\delta t=\frac{0.1}{\max\left( \theta,2\mu,2\gamma,2\upsilon,2\zeta\right)}$$

The exponential growth and the ongoing (de)methylation of the cancer cell population are treated in two steps, first the population of the cancer grows according to the deterministic exponential growth equation (rounded to the closest integer):

$$N_{e}\left( t \right)=e^{\theta\left( t-\tau\right)}$$

With the new cells assigned to each population according to the multinomial distribution:

$$\left( m_{g}, k_{g}, w_{g} \right)\sim\mathrm{Multinomial}\left( N_{e}\left( t+\delta t \right)-N_{e}\left( t \right), \left( \frac{m\left( t \right)}{N_{e}\left( t \right)},\frac{k\left( t \right)}{N_{e}\left( t \right)},\frac{w\left( t \right)}{N_{e}\left( t \right)} \right) \right)$$

$$\left( m^{'}\left( t \right), k^{'}\left( t \right), w^{'}\left( t \right) \right)=\left( {m\left( t \right)+m}_{g},k\left( t \right)+ k_{g},w\left( t \right)+ w_{g} \right)$$

The flow from the homozygous methylated to the heterozygous methylated population due to ongoing demethylation can then be calculated as:

$$\delta_{m\to k;t}\sim\mathrm{Binomial}\left( m^{'}\left( t \right),2\gamma\delta t \right)$$

Similarly, the flow from the homozygous unmethylated to the heterozygous methylated population is:

$$\delta_{w\to k;t}\sim\mathrm{Binomial}\left( w^{'}\left( t \right),2\mu\delta t \right)$$

To calculate the flow from the heterozygous population to the homozygous unmethylated and methylated populations, we first calculate the total number of cells that change state and then work out whether they flow to the homozygous methylated or unmethylated population:

$$\delta k_{t}\sim\mathrm{Binomial}\left( k^{'}\left( t \right),\left( \upsilon+\zeta\right)\delta t \right)$$

$$\delta_{k\to m;t}\sim\mathrm{Binomial}\left( \delta k_{t},\frac{\upsilon}{\upsilon+\zeta} \right)$$

$$\delta_{k\to w;t}=\delta k_{t}-\delta_{k\to m;t}$$

Updating the system at time $t + \delta t$ is then:

$$m\left( t+\delta t \right)=m^{'}\left( t \right)+\delta_{k\to m;t}-\delta_{m\to k;t}$$

$$k\left( t+\delta t \right)=k^{'}\left( t \right)+\delta_{k\to m;t}+\delta_{w\to k;t}-\delta k_{t}$$

$$w\left( t+\delta t \right)=w^{'}\left( t \right)+\delta_{k\to w;t}-\delta_{w\to k;t}$$

Note that the reason for this formulation, rather than simply calculating the number of cells that have switched from one state to another via each process (e.g. following a Poisson process) separately, is to strictly ensure that the total cell number is conserved and each population is strictly positive. The methylation fraction at time $t$ can be simply calculated as:

$$\beta_{c}\left( t \right)=\frac{k\left( t \right)+2m\left( t \right)}{2N_{e}\left( t \right)}$$

The above assumes a perfectly pure tumour sample, however, there will inevitably be a degree of contamination from normal cells. To account for this contamination, we assumed that the contaminating normal cells did not share a recent common ancestor, so the measured methylation of a tumour sample with purity $\rho$ would be the weighted average of the cancer methylation at that locus, $\beta_{c}\left( t \right)$, and the average methylation of the contamination normal cells, $\Lambda$:

$$\beta\left( t \right)=\rho\beta_{c}\left( t \right)+\left( 1-\rho\right)\Lambda$$

This assumption was informed by the observation that the standard deviation of the different cancer sample’s fCpG methylation distributions correlated linearly with the tumour purity (fig. S14E).

Using the recursive relationship above, a single stochastic path can be generated. The methylation fraction probability distribution can then be numerically estimated by generating a series of $n_{sim}$ independent samples from the above process, the distribution of which will approximate the underlying hidden Markov model distribution.

### The Likelihood Function for a Growing Population

The stochastic model above allowed samples to be drawn from the distribution of fCpG methylation values given a set of parameters $\left\{ \theta,\tau,\mu,\gamma,\upsilon,\zeta,\rho,\beta_{n} \right\}$. We can use these samples to form an estimate of the likelihood function for a given set of data, $\overset{⃑}{y}$. First, we can bin the $n_{sim}$ stochastically generated $\beta\left( t \right)$ into $J$ bins with width $\frac{1}{J}$, to estimate a probability mass function (p.m.f) $P\left( \beta_{j}|\theta,\tau,\mu,\gamma,\upsilon,\zeta,\Lambda\right)$ for bins with centres $\beta_{j}=\frac{2j-1}{2B}$ ($j=1, 2, \ldots, J$).

To account for the noise introduced by the methylation array, we follow a similar approach as in (Gabbutt et al. 2022), first employing a linear transform to account for the background noise of the array: $x_{j} = (\varepsilon- \Delta)\beta_{j} + \Delta$. Then, we can model the measured methylation fraction values $y_{i}$ as a mixture of beta distributed random variables, drawn from a beta distribution with mean $x_{j}$ and precision $\kappa$, weighted by the p.m.f. $P\left( \beta_{j}|\theta,\tau,\mu,\gamma,\upsilon,\zeta,\Lambda\right)$. Assuming that each measured fCpG site is independent, such that the likelihood of a set of fCpG loci is the product the per-fCpG likelihood, the likelihood is then:

$$P\left( y_{i}|\beta_{j},\Delta,\varepsilon,\kappa\right)=\frac{{y_{i}}^{\kappa x_{j}-1}\left( 1-y_{i} \right)^{\kappa\left( 1-x_{j} \right)}-1}{B\left( \kappa x_{j},\kappa\left( 1-x_{j} \right) \right)}$$

$$\mathcal{L}\left( \theta,\tau,\mu,\gamma,\upsilon,\zeta,\beta_{n},\Delta,\varepsilon,\kappa|\overset{⃑}{y} \right)=\prod_{i=1}^{n_{sim}} \sum_{j=1}^{J} P\left( y_{i}|\beta_{j},\Delta,\varepsilon,\kappa\right)P\left( \beta_{j}|\theta,\tau,\mu,\gamma,\upsilon,\zeta,\Lambda\right)$$

It is often convenient to work with the log-likelihood, rather than the likelihood directly. Employing the logsumexp function, $LSE\left( x_{1},\ldots, x_{n} \right)=\log\left( e^{x_{1}}+,\ldots, +e^{x_{n}} \right)$, the log-likelihood is:

$$\log\left( \mathcal{L}\left( \theta,\tau,\mu,\gamma,\upsilon,\zeta,\beta_{n},\Delta,\varepsilon,\kappa|\overset{⃑}{y} \right) \right)=\sum_{i=1}^{N} LSE\left( \log\left( P\left( y_{i}|\beta_{j},\Delta,\varepsilon,\kappa\right) \right)+\log\left( P\left( \beta_{j}|\theta,\tau,\mu,\gamma,\upsilon,\zeta,\Lambda\right) \right) \right)$$

Here, we have modelled the growth rate and the time at which the MRCA emerged as independent; however, as we are assuming exponential growth, if we vary these two parameters independently, we implicitly assume that the total cancer burden can take on implausibly high values. To penalise combinations of $\theta$ and $\tau$ that yield such high values, we add a penalisation factor for cancer burdens outside the plausible range $\left\{ {N_{e}}^{min},{N_{e}}^{max} \right\}$.

$$\mathcal{L}_{penalise}= e^{-\frac{{N_{e}}^{min}}{N_{e}}}-e^{-\frac{{N_{e}}^{max}}{N_{e}}}$$

Note that this is equivalent to placing a joint dependent prior over $\theta$ and $\tau$.

### Stochastic likelihood calculation correction

The log-likelihood calculation above differs from a classical likelihood calculation in that $\log\left( \mathcal{L} \right)$ is not an exact value, but rather an approximation of the “true” log-likelihood which should become more exact as $n_{sim}\to\infty$. To test whether this estimator is systematically biased, we simulated a set of $\overset{⃑}{y}$ data with a set of known parameters, then calculated the $\log\left( \mathcal{L} \right)$ with the same set of parameters for a range of $n_{sim}$ values (fig. S15A).

We find that at low $n_{sim}$ values the approximation tends to systematically underestimate the log-likelihood, whilst as $n_{sim}$ increases, the calculated log-likelihood tends to asymptote to a single value. The increase in the mean estimated log-likelihood with $n_{sim}$ can be rationalised if one considers that we are attempting to estimate a continuous pdf by sampling from it, and as $n_{sim}$ increases, the stochastic noise from sampling is reduced and the approximation improves. In this sense, the fit at the peak of the log-likelihood distribution truly is more accurate, on average, as $n_{sim}$ increases, hence the calculated log-likelihood is higher. Unsurprisingly, the standard deviation of the calculated log-likelihood values falls as the number of simulated paths increases.

We found that the bias associated with having a finite $n_{sim}$ varied as $\log\left( \mathcal{L}_{bias} \right)\sim\frac{1}{n_{sim}}$. By varying different parameters, we found the log-likelihood bias could be estimated as:

$$\log\left( \mathcal{L}_{bias} \right)\approx-\frac{0.5+0.005\kappa^{\frac{4}{3}}}{n_{sim}}$$

Correcting for the bias yields log-likelihood estimates with a mean that does not vary with $n_{sim}$ (fig. S15B). Hence, the bias-corrected cancer burden penalised log-likelihood is:

$${\log\left( \mathcal{L} \right)}^{'}=\log\left( \mathcal{L} \right)+\log\left( \mathcal{L}_{penalise} \right)-\log\left( \mathcal{L}_{bias} \right)$$
